# The Humanitarian-Development Nexus and Sexual and Reproductive Health Interventions in Fragile Settings: A Scoping Review

**DOI:** 10.1101/2024.10.14.24315444

**Authors:** Amany Qaddour, Hannah Tappis, Stefany Lazieh, Ava Ward, Paul Spiegel

## Abstract

The concurrent need for both humanitarian and development assistance in fragile settings and protracted emergencies has been termed the humanitarian-development nexus (HDN) or the humanitarian-development-peace nexus (HDpN). We undertook a scoping review on the operationalization of the HDpN for sexual, reproductive, maternal, newborn, child, and adolescent health interventions in fragile settings. We screened 2,183 publications, of which 29 peer-reviewed and 16 grey literature publications met inclusion criteria. No included studies focused on peace aspects within the HDpN and very few focused on child and adolescent health. Publications by humanitarian authors often classified maternal and newborn health as a component of sexual and reproductive health (SRH). Data extraction and analysis focused on three overarching themes: SRH prioritization across the HDN, the transition between minimum and comprehensive services, and health systems strengthening. This review provides concrete guidance on how to operationalize the HDN for SRH interventions in fragile settings. Expansion of SRH preparedness measures is necessary given the current trajectory of the climate emergency and other destabilizing events. The ability to flexibly transition between minimum and comprehensive services is important for maintaining service continuity in crisis-affected settings. COVID-19 proved to be a significant disruptor of SRH services, and a key inflection point in the collaboration between humanitarian and development practitioners. The use of task-shifting, decentralization, and telemedicine were approaches that may be adopted to maintain service delivery according to different contexts. Lastly, strengthening health systems was identified as essential across the HDN. With more crises extending for years, the wider literature has emphasized the necessity of health systems strengthening for reaching the Sustainable Development Goals, including in fragile settings.

## INTRODUCTION

The overlap and intersection of humanitarian and development assistance in complex settings has been termed the humanitarian-development nexus (HDN), which focuses on the need to address the vulnerability of people before, during, and after crisis (1). The humanitarian-development-peace nexus (HDPN/HDpN) or “triple nexus” expands the concept one step further to incorporate peace efforts, the latter traditionally ascribed to actors with peacekeeping and peacebuilding mandates (2). Efforts to address root causes of conflict and other underlying drivers of fragility have demonstrated the complexities of integrating peace aspects into the HDN given that these often extend beyond the parameters of traditional humanitarian and development assistance mandates.

While nexus terminology continues to evolve and various frameworks have emerged over the last 40 years, it is apparent that the traditional binary designation of assistance as either humanitarian or development is no longer appropriate, as we continue to observe an increase in fragility across many countries and regions, globally (3–6). Some countries once deemed relatively stable have slipped into deep instability (e.g., Sudan, Ukraine, Venezuela), whereas other countries continued to be plagued by volatility and fragility (e.g., Yemen, Myanmar, Somalia) (7,8). Conflicts have also become more protracted, violent, and complex in nature, often spanning years and decades, with many recently (and aptly) termed the “new forever wars” by some scholars (9–11). Political instability and stalled peace processes remain hallmark features in many of these settings, and we can expect that political upheaval may perpetuate fragility in different countries given there are a record 64 elections around the world in 2024, with 43 (nearly 70%) of these occurring in low- and middle-income countries (LMICs) (12,13). Changes in the political landscape of these countries will inevitably have an impact on agenda-setting for the health sector, such as sexual and reproductive health and rights, (e.g., the “politics of SRHR” (14)), or the inclusion of refugees in national health plans in the case of refugee-hosting countries (15).

Other recurrent shocks and stresses have contributed to the heightened fragility in many LMICs, including factors of economic insecurity, mass displacement, gender inequality, and major infectious disease outbreaks, such as the COVID-19 pandemic, which pushed nearly 150 million people into extreme poverty (16). In addition to these factors, climate emergencies will continue to have catastrophic ramifications, with estimates of 14.5 million deaths and 1.2 billion displacements by 2050 (17–19). These issues have overlapped and further compounded the vulnerability of populations. In an effort to address such vulnerability, different measures have emerged to mitigate the impact of shocks and stresses in fragile settings, including disaster risk reduction (DRR, e.g., the Sendai Framework) (20), preparedness for climate emergencies (e.g., anticipatory action) (21,22), and efforts to strengthen health systems and build resilience (23,24). Despite these measures, fragility in LMIC settings continues to increase. As of 2022, over 1.9 billion people lived in fragile contexts (8). Over 117.3 million people were forcibly displaced in 2023, including the stateless, internally displaced persons (IDPs), and refugees (25,26). The majority (75%) of refugees in 2023 were hosted in LMICs, further constraining the resources available in these settings (26,27). Global figures have also shown that refugees are displaced outside of their country of origin for an average of 20 years and IDPs for over 10, warranting a more pronounced focus on rights-based, durable solutions for displaced communities (28,29).

The sustainability of services in fragile settings has proven challenging given the difficulties in balancing interventions of a short-term (i.e., humanitarian) and medium- or long-term (i.e., development) nature. It has become clear that in settings routinely affected by recurrent shocks and stressors, it is often necessary to deliver humanitarian and development assistance simultaneously. This phenomenon has become increasingly more common as countries with significant humanitarian needs in one geographic area that may also be primed for early recovery or development assistance in other locations. Relapses in fragility and the transition from an acute to post-crisis phase is no longer a linear process, and service design and delivery reforms in fragile settings have proven necessary. Such reforms are also rooted in other aspects of the aid sector, including more recent demands to address structural inequalities, racism, localization commitments, decolonization, and power imbalances between Global North and Global South countries (30–33).

Within fragile settings, women and children are particularly at risk, as they make up over 75% of those in need and are disproportionately affected by crisis (11,34). Health service delivery in these settings is extremely challenging given significant health workforce shortages, which estimated to increase dramatically by 2030 (35). In addition, violence toward health care workers in conflict settings and attacks to health infrastructure remain pervasive: approximately 487 health workers were killed, 445 arrested, and 240 kidnapped in 2023 (36). The delivery of sexual, reproductive, maternal, newborn, child, and adolescent health (SRMNCAH) services is life-saving and critical for reducing rates of mortality and morbidity (37–39). For this reason, and regardless of the context or setting, there should be a commitment to ensure the needs of women and children are taken into account in fragile settings across all forms of assistance delivered (e.g., humanitarian, development). Various strategies and tools guide this massive undertaking, but practical examples are still needed to navigate the complexities and evolving nature of service delivery in fragile settings.

### Objective of Review

The objective of this review was to map existing literature related to the HDpN and SRMNCAH interventions in fragile settings. We sought to understand how HDpN concepts are described and summarized the reported successes and challenges in applying the nexus concepts to SRMNCAH interventions in fragile settings. It builds on the report “The Humanitarian-Development Nexus: A Framework for Maternal, Newborn, and Child Health, Voluntary Family Planning, and Reproductive Health in Fragile Settings,” as developed by Johns Hopkins Center for Humanitarian Health in partnership with MOMENTUM Integrated Health Resilience (40). As a precursor to the scoping review, the landscape analysis examined the HDN and its application to health interventions along with the development of a conceptual framework outlining the key concepts, core components, and cross-cutting themes that were relevant to these interventions.

## METHODS

We conducted a review of peer-reviewed publications and grey literature using the Prisma Extension for Scoping Reviews (PRISMA-ScR) and the methodological framework for scoping reviews of Levac et. al, consisting of the five-stage process where we set out to identify the research question(s); identify relevant studies; determine search criteria; chart the data; and collate, synthesize, and report the results (41,42). To the best of our knowledge, a scoping review of the HDpN and its application to SRMNCAH does not exist in the literature, and a comprehensive synthesis of this topic has not yet been conducted.

### Protocol and Registration

The title and protocol for this review was formally registered in the Open Science Framework on October 8, 2022 [DOI: https://doi.org/10.17605/OSF.IO/6BSE4].

### Search Strategy

The databases searched for peer-reviewed literature included Web of Science, Scopus, PubMed, and CINAHL. Forward citation tracking was conducted. Grey literature sources were also included in the review given that a sizable portion of humanitarian and development program guidance and practices are documented through bilateral/multilateral organizations and non-governmental organizations (NGOs). Using relevant databases and libraries, the PRISMA checklist guided and structured this review (see **S1 File**). The overall search strategy was continuously refined throughout this review to ensure new themes and keywords were added to the search strategy as they emerged in the literature through the period of March 31, 2023. Final search terms are summarized in **Table 1** with the search strategy for each database for peer-reviewed literature provided in the supporting information section (**S2 File**). Covidence software was used to manage the search and screening process for peer-reviewed publications. Title/abstract screening was conducted by two independent reviewers to produce a short list of articles, followed by full text screening by dual review for agreed upon articles to confirm the articles met eligibility criteria. A third reviewer reviewed any areas of discordance for consensus. A manual, online search for grey literature was conducted through various humanitarian and development resource hubs, summarized in the supporting information section (**S3 File**). These publications were organized using Microsoft Excel. **Figure 1** outlines the literature search processes as per PRISMA statement.

**Figure 1.**
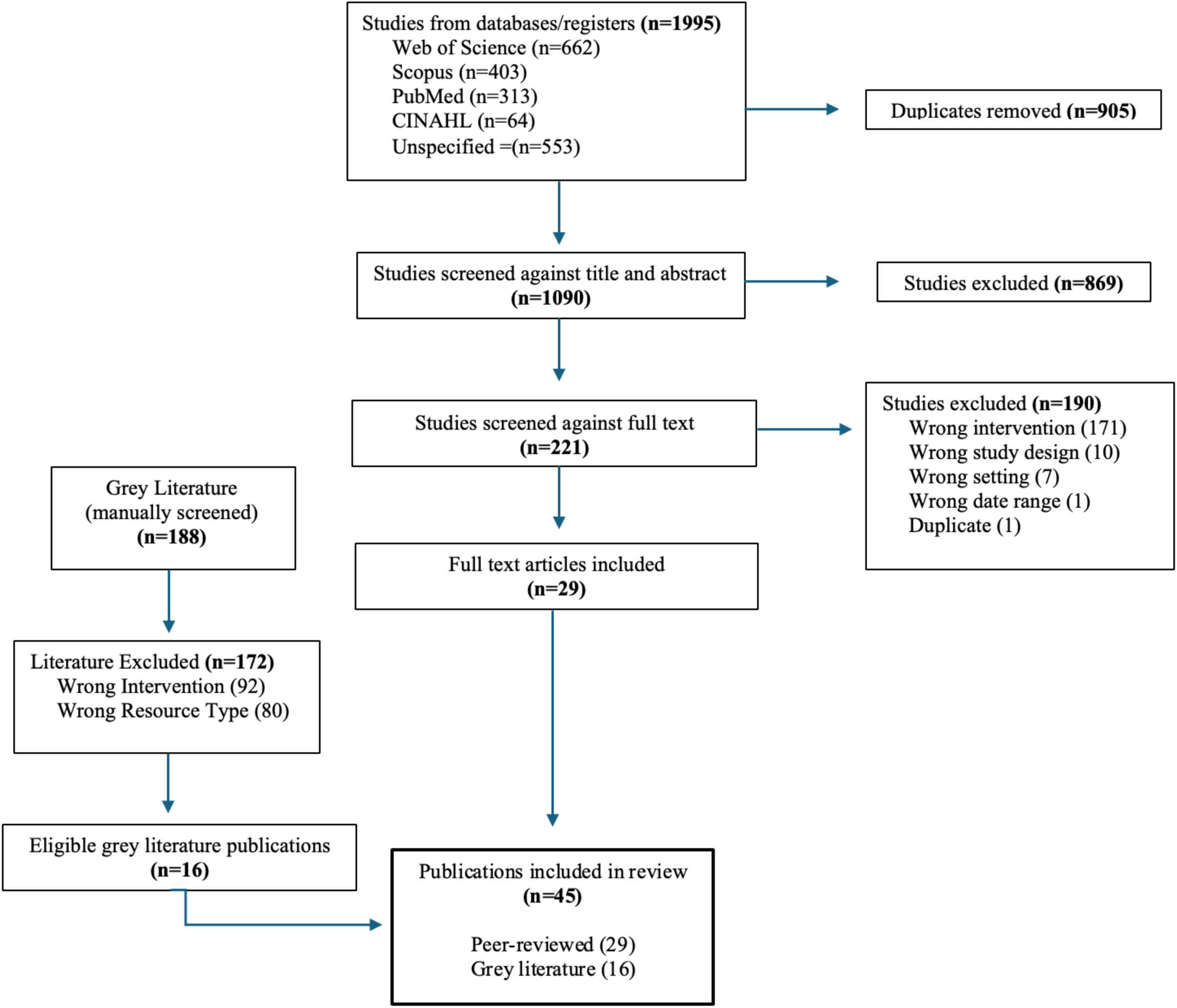
**Literature Search**

**Table 1.**
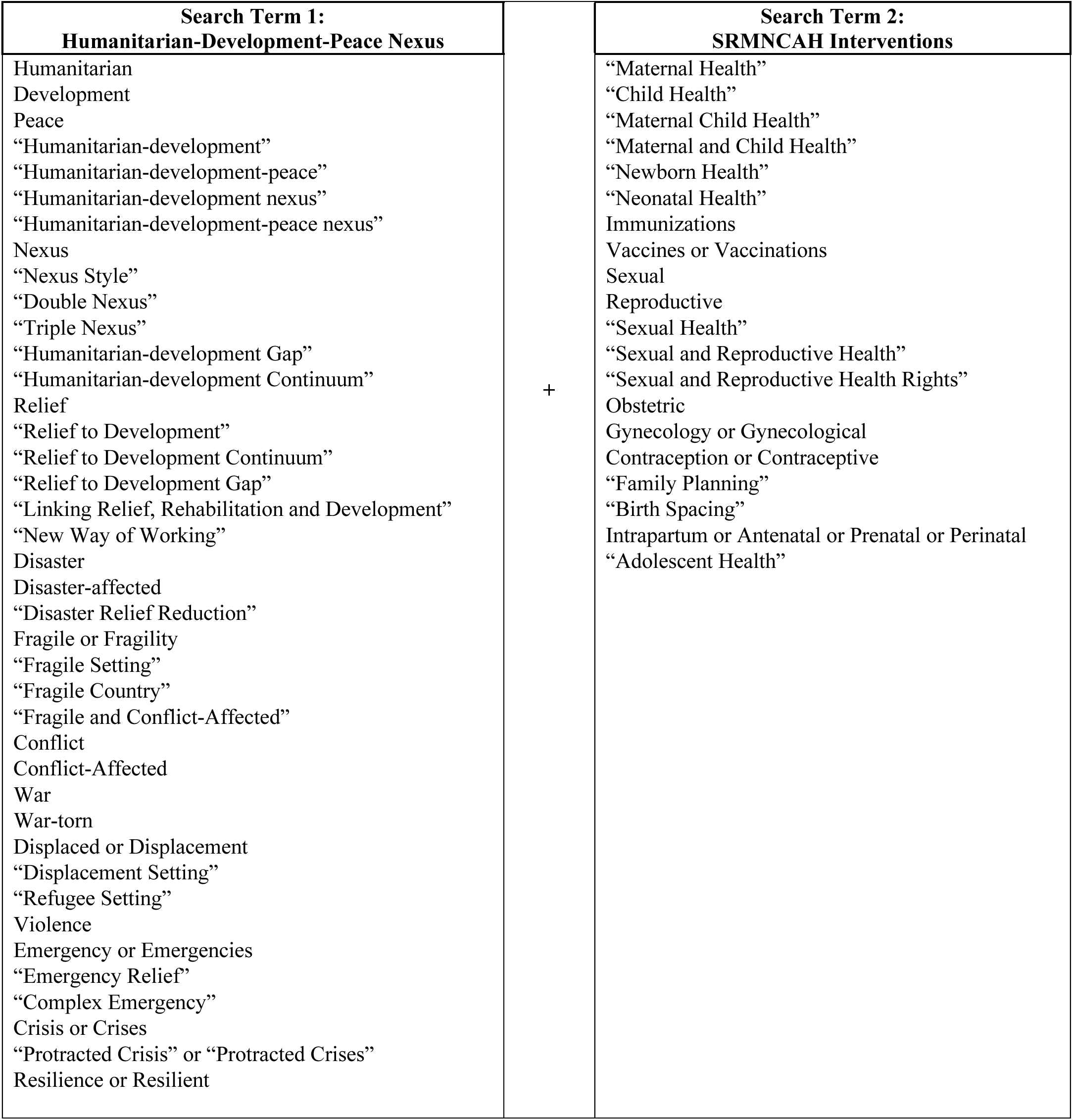
Search Terms.

### Eligibility Criteria

The screening criteria included relevant literature published with a focus on the HDpN and SRMNCAH interventions in fragile settings. Given the broad definition and limitations in ascribing contexts as humanitarian, development, and/or fragile, the search included all LMICs, as per World Bank designation, which also met one or more of the following criteria from for the period of 1990-2023 (43–46):

▪ Launch of a Humanitarian Response Plan and/or Flash Appeal
▪ IASC Humanitarian System-Wide Emergency Activation or Active Scale Up:

∘Emergency Activation (L3 Response) (*2012–2018*)
∘Active Scale Up *(2018-Present)*
▪ World Bank Fragility Designation:

∘ Low Income Category Under Stress (LICUS) (*2004–2008*)
∘ Fragile States List *(2009-2010)*
∘ Harmonized List of Fragile Situations *(2011-2020)*
∘ Fragile and Conflict-Affected Situations (FCS) *(2020-Present)*

Territories that were not included in LMIC listings but still met the criteria above were included in this review (e.g., Chechnya, Occupied Palestinian Territories). Within countries that met eligibility criteria, affected populations included those from host communities, IDPs, and refugees. Resettled refugees and asylum seekers in high-income countries were not included. A summary of countries organized by LMIC status and relevant fragility designation is available in the supporting information section (**S4 File**). SRMNCAH interventions were included and interventions of other sectors were excluded (e.g., food security, nutrition, education). The search covered literature published between January 1, 1990, through March 31, 2023. Only publications in English were included in the search, and all other languages were excluded. Eligibility criteria are summarized in **Table 2**.

**Table 2.**
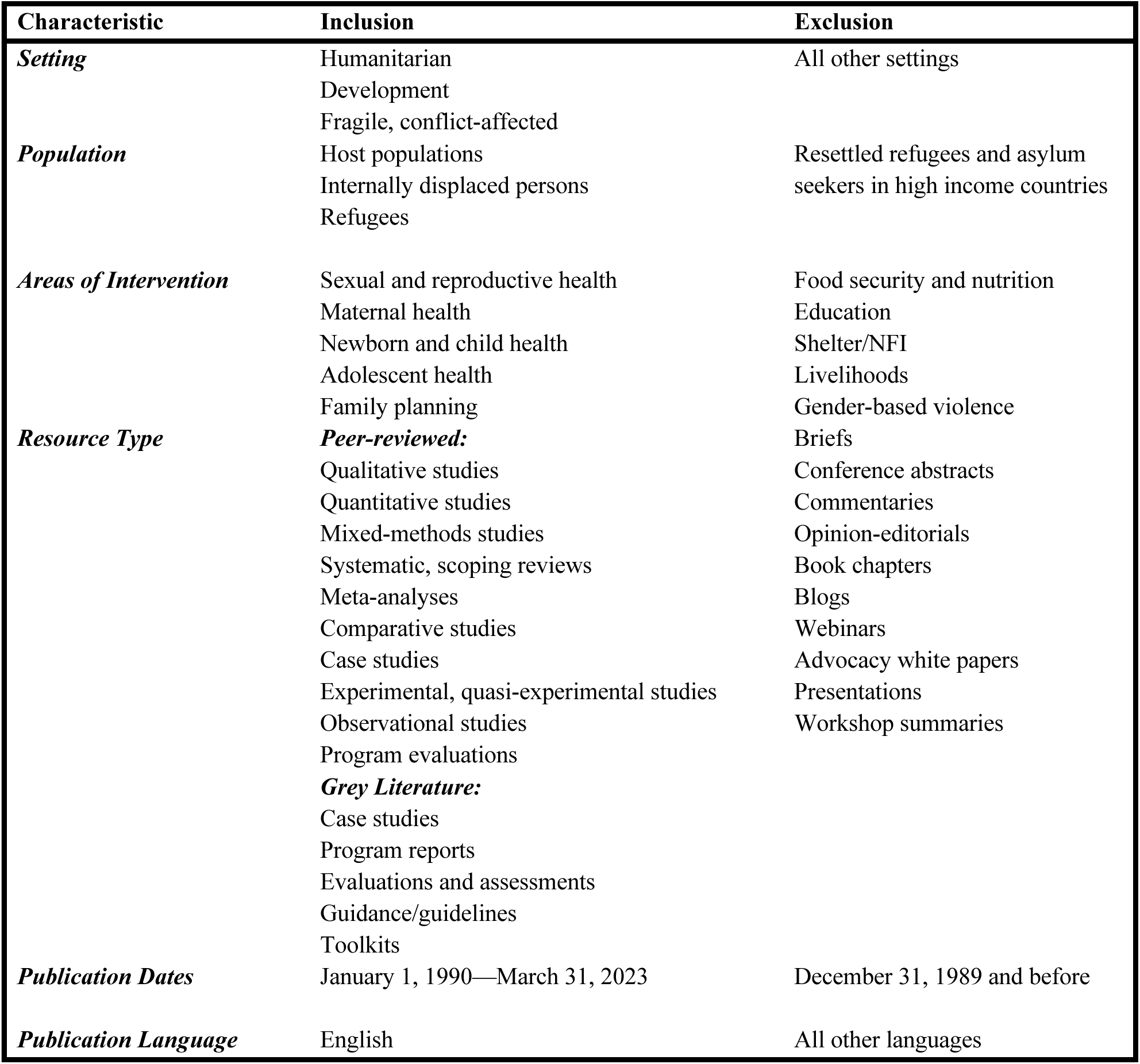
Eligibility Criteria.

### Data Extraction and Analysis

Selected literature that met the criteria for this review was charted and organized by geographic context (country/region), area of intervention within SRMNCAH, and thematic area(s) of focus or significance as it may relate to the HDpN. Although the search strategy did not focus on nutrition interventions, several of the publications included nutrition as integrated within or alongside maternal, newborn, child, and adolescent health (MNCAH) interventions. The information was organized within the data extraction tool developed. Content analysis of the extracted data took place and allowed for the identification of key themes and findings from the literature. Coding of relevant data was independently taken by a member of our research team along with review by a second member for concurrence.

Thematic areas were pre-determined (*a priori*) and guided by the HDN landscape analysis and conceptual framework previously developed by the research team, which also drew from multiple frameworks and theories of change, including the HDN Conceptual Framework, WHO Health Systems Framework, and UN Sendai Framework for Disaster Risk Reduction (20,40,47). Themes were continuously refined and re-organized throughout the analysis phase, as the process was iterative. Thematic areas were intended to draw out nexus-relevant content, with the caveat that publications did not always use explicit nexus terminology. There were often multiple, overlapping themes covered within one publication or instances where authors had published both a peer-reviewed and grey literature version on the same topic. In both instances, findings were summarized once to avoid repetition. Additionally, thematic findings were only reported in instances where substantive information was presented, with a prioritization of major themes covered in each respective publication.

## RESULTS

### Characteristics of Included Papers

We identified 45 publications on the HDpN and its operationalization for SRMNCAH interventions in fragile settings. For humanitarian actors, sexual and reproductive health (SRH) programming included maternal and newborn health (MNH), family planning (FP), gender-based violence (GBV) prevention and response, and management of sexually transmitted infections (STIs), including HIV. SRH was organized and clustered differently by development actors and was often organized as MNH, FP/RH, HIV, GBV, MNCAH, RMNCAH, or SRMNCAH, among other classifications. Of the total publications, 29 were peer-reviewed and 16 were grey literature publications. Publications encompassed 39 different countries across 5 different regions, including Africa (*n=21*), Asia and Pacific (*n=12*), Middle East (*n=4*), Latin America (*n=1*), and Europe (*n=1*), as summarized in **Table 3**. While our search strategy encompassed HDpN and SRMNCAH, the bulk of the relevant literature focused on the HDN and SRH. There were none that focused on peace aspects within the HDpN and very few focused on child and adolescent health. Content on these areas addressed service coverage but not themes related to the HDN. For this reason, results will focus on SRH as used in the humanitarian sector. The majority of publications focused on conflict and/or post-conflict settings (*n=36*), followed by epidemics and outbreaks (*n=14*), and climate shocks and natural disasters (*n=8*). **Table 4** shows the number of publications reporting on contexts and themes of interest. Note that the totals exceed the number of publications, as there was an overlap in the interventions, contexts, and thematic areas covered in some of the literature. Findings were organized with a focus on the most significant themes in each publication. **Table 5** provides an expanded summary of all results included in this review, organized by author, year, country, area of intervention, context, and thematic area.

**Table 3.**
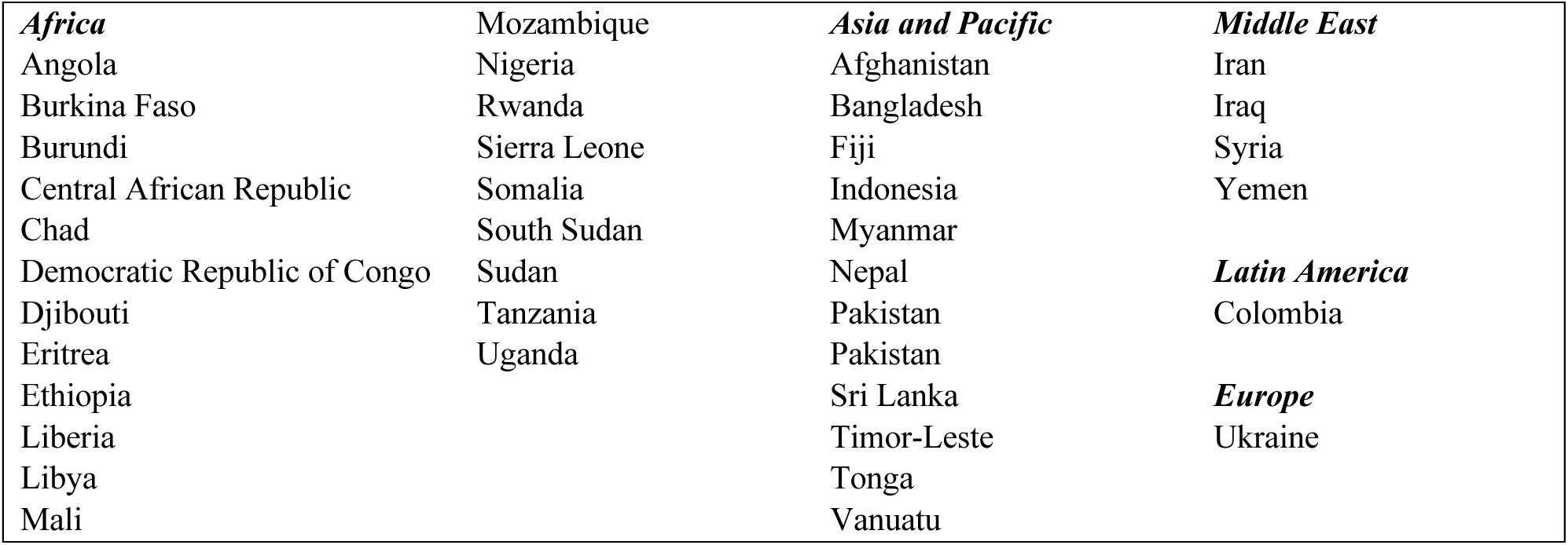
List of Countries included in Scoping Review, Organized by Region.

|  |  |  |  |
| --- | --- | --- | --- |
| ***Africa*** | Mozambique | ***Asia and Pacific*** | ***Middle East*** |
| Angola | Nigeria | Afghanistan | Iran |
| Burkina Faso | Rwanda | Bangladesh | Iraq |
| Burundi | Sierra Leone | Fiji | Syria |
| Central African Republic | Somalia | Indonesia | Yemen |
| Chad | South Sudan | Myanmar |  |
| Democratic Republic of Congo | Sudan | Nepal | ***Latin America*** |
| Djibouti | Tanzania | Pakistan | Colombia |
| Eritrea | Uganda | Pakistan |  |
| Ethiopia |  | Sri Lanka | ***Europe*** |
| Liberia |  | Timor-Leste | Ukraine |
| Libya |  | Tonga |  |
| Mali |  | Vanuatu |  |

**Table 4.** Scoping Review Results at a Glance.

| **Intervention** | **No. of Articles (n)** |
| --- | --- |
| SRH | 34 |
| MNCAH | 20 |
| **Context** |  |
| Climate Shocks and Natural Disasters | 8 |
| Conflict, Post-conflict | 36 |
| COVID-19, epidemics, outbreaks | 14 |
| **Themes & Sub-themes** |  |
| Theme 1. SRH Prioritization Across the HDN | 10 |
| *Sub-theme 1.1. Integrating SRH within DRR and EPP* | 12 |
| *Sub-theme 1.2. Exit Strategies in Recovery Scenarios* | 4 |
| Theme 2: Transition between Minimum and Comprehensive Services | 13 |
| *Sub-theme 2.1: Adaptations during the COVID-19 Pandemic and other Outbreaks* | 12 |
| Theme 3: Health Systems Strengthening | 12 |
| *Sub-theme 3.1. Health Systems Strengthening through Workforce Development* | 17 |
| *Sub-theme 3.2. Other Health System Strengthening Reforms* | 19 |

**Table 5.**
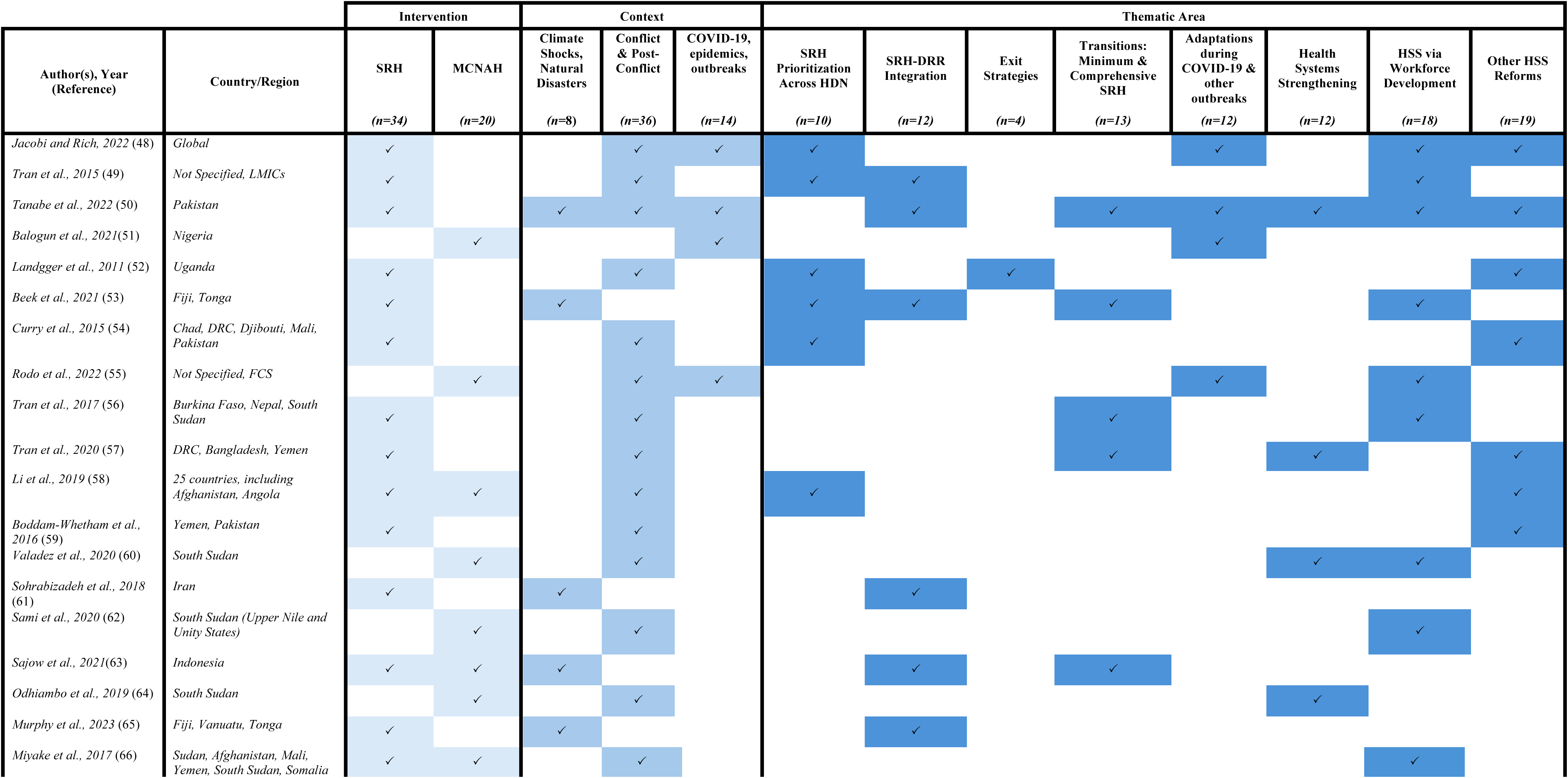
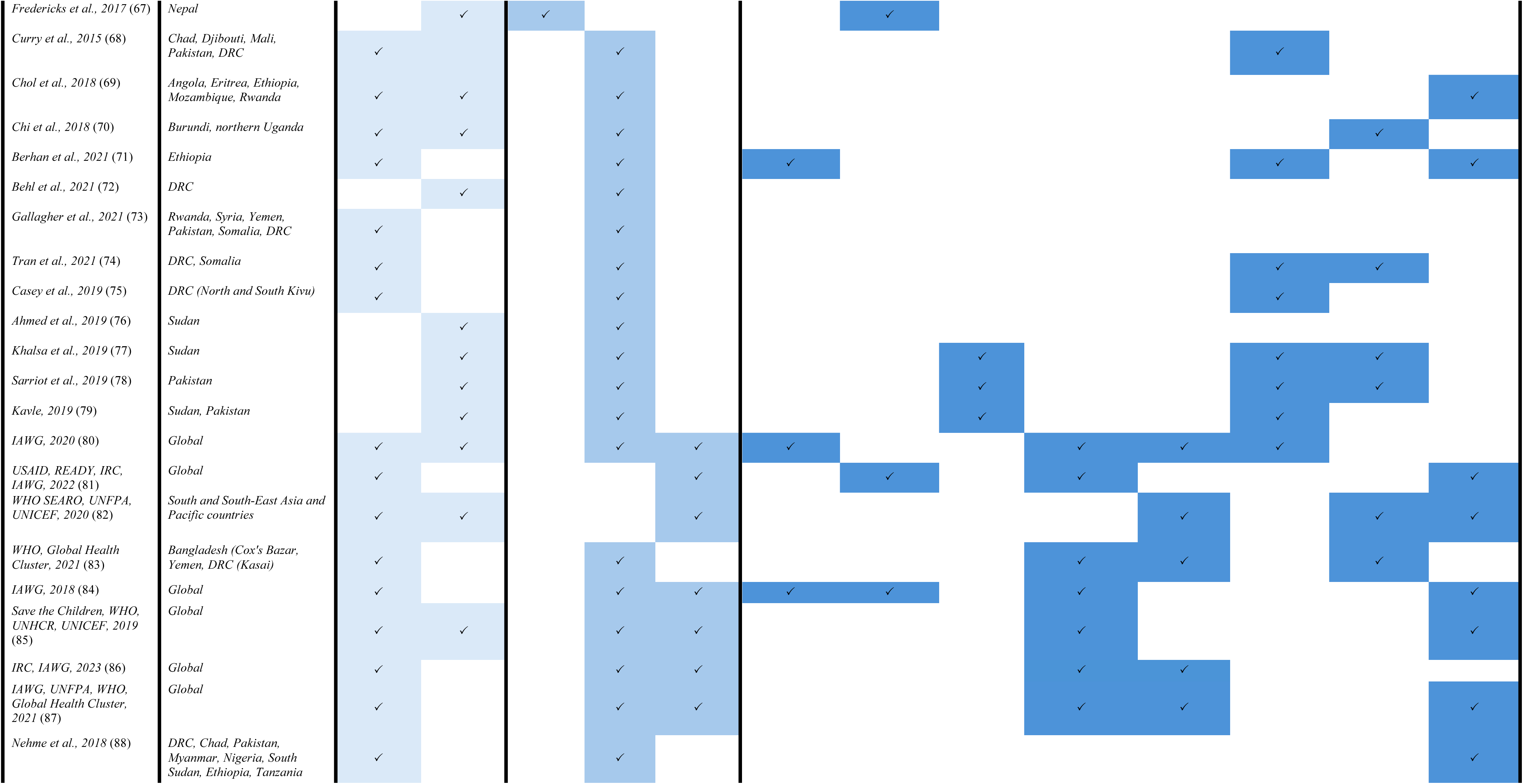
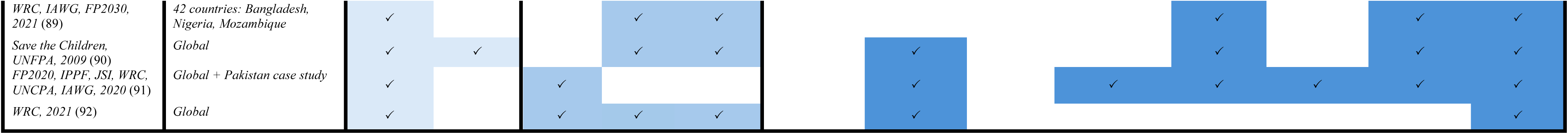
Summary of Included Studies by Intervention Type, Context, and Themes Relevant to the HDN.

### Theme 1. SRH Prioritization Across the Humanitarian-Development Nexus

The literature highlighted varied perspectives on SRH prioritization across the humanitarian-development nexus. A total of 10 publications discussed prioritization, including seven on SRH (49,52,53,58,71,80,84) and three with a narrower focus on FP (48,54,89). SRH funding from development sources (ODA) in conflict-affected countries (with simultaneous humanitarian funding) demonstrated a tremendous increase in funds (298% between 2002-2011) (54) amidst a broader increase in development assistance (seven-fold between 2003-2017) in these countries (58). However, it was noted that the majority of these SRH funds were channeled to HIV/AIDS, not other services within the scope of SRH (54,58). Prioritization of other health services over SRH was emphasized in different publications, with perspectives of SRH as “buried within the health sector,” in northern Uganda during its transition from the health cluster to sector working groups (52); being “lost within the health cluster” in the case of Fiji’s cyclone emergency response among other health and non-health needs (e.g., food, shelter) (53); or less prioritized amidst competing health demands during COVID-19 and other infectious disease outbreaks (discussed in sections to follow) (80). In Ethiopia’s journey to attain universal health coverage (UHC) and efforts to integrate SRH, clinical services for pregnant women and newborns and FP services were delivered through the UHC package given they aligned with the Millenium Development Goals, while other sexual health services were not consistently offered for the same reason (71).

Divergent perspectives were outlined in the literature in terms of whether SRH and FP services were recognized as components of humanitarian or development assistance, and subsequently, whether they were the responsibility of humanitarian or development actors (48,49,54,58,89). The Inter-Agency Working Group on Reproductive Health in Crises (IAWG)’s ten year review of RH in humanitarian settings stressed that RH services should fall under the purview of both forms of assistance given the “emergency management cycle” which necessitates both types of actors be involved (49), while also documenting this shift in its Inter-Agency Field Manual given the nature of more recent crises, including those of a protracted nature or “non-linear trajectory” (80). Differing perspectives on FP as a life-saving activity was discussed across multiple fragile contexts (48,54,89), along with recommendations for consensus building around this notion (89).

### Sub-Theme 1.1. Integrating SRH within Disaster Risk Reduction

The need to integrate SRH within disaster risk reduction (DRR) frameworks and emergency preparedness plans (EPP) was a recurrent theme across the literature given, building on the premise of SRH prioritization and readiness to deliver and ensure continuity of SRH services during periods of crisis. The institutionalization of SRH within disaster risk management and national development plans was highlighted as a critical component for formalized SRH-DRR integration. This integration was contingent upon the prioritization of RH within the health system, readiness to implement the Minimum Initial Service Package (MISP) for SRH in crisis settings, and measures to introduce or re-integrate comprehensive SRH services in the aftermath. These measures essentially guide the implementation of DRR to mitigate the severity of crisis, while EPP can help enhance effectiveness of a response and accelerate the transition from an emergency to post-emergency scenario, with the caveat that this is not always a linear process. This transition was defined as the “continuum of an emergency,” in the Inter-Agency Field Manual for RH in Humanitarian Settings with visual incorporation of this concept and the inclusion of **Figure 2** below in six of the publications (57,81,84,86,88,90). A total of 12 publications discussed the integration of SRH within DRR (49,50,53,61,63,65,67,81,84,90–92). Of these articles, **six** were peer-reviewed and documented emergency response to a natural disaster or climate shock, including floods in Pakistan (50), earthquakes in Iran and Nepal (61,67), volcanic eruption in Indonesia (63), cyclones in Fiji and Tonga (53), and more broadly for climate-related disasters across the Pacific region, including Fiji, Tonga, and Vanuatu (65). Generalized global guidance for SRH continuity was outlined in the IAWG’s Inter-Agency Field Manual for RH in Humanitarian Settings, which has served as the basis for other guidance and implementation efforts, including the READY Initiative’s preparedness and response checklist for outbreaks using MISP objectives (81,84). Other guidance focused on the involvement of communities (50,92) and adolescents and youth (65,91) in SRH preparedness efforts.

**Figure 2.**
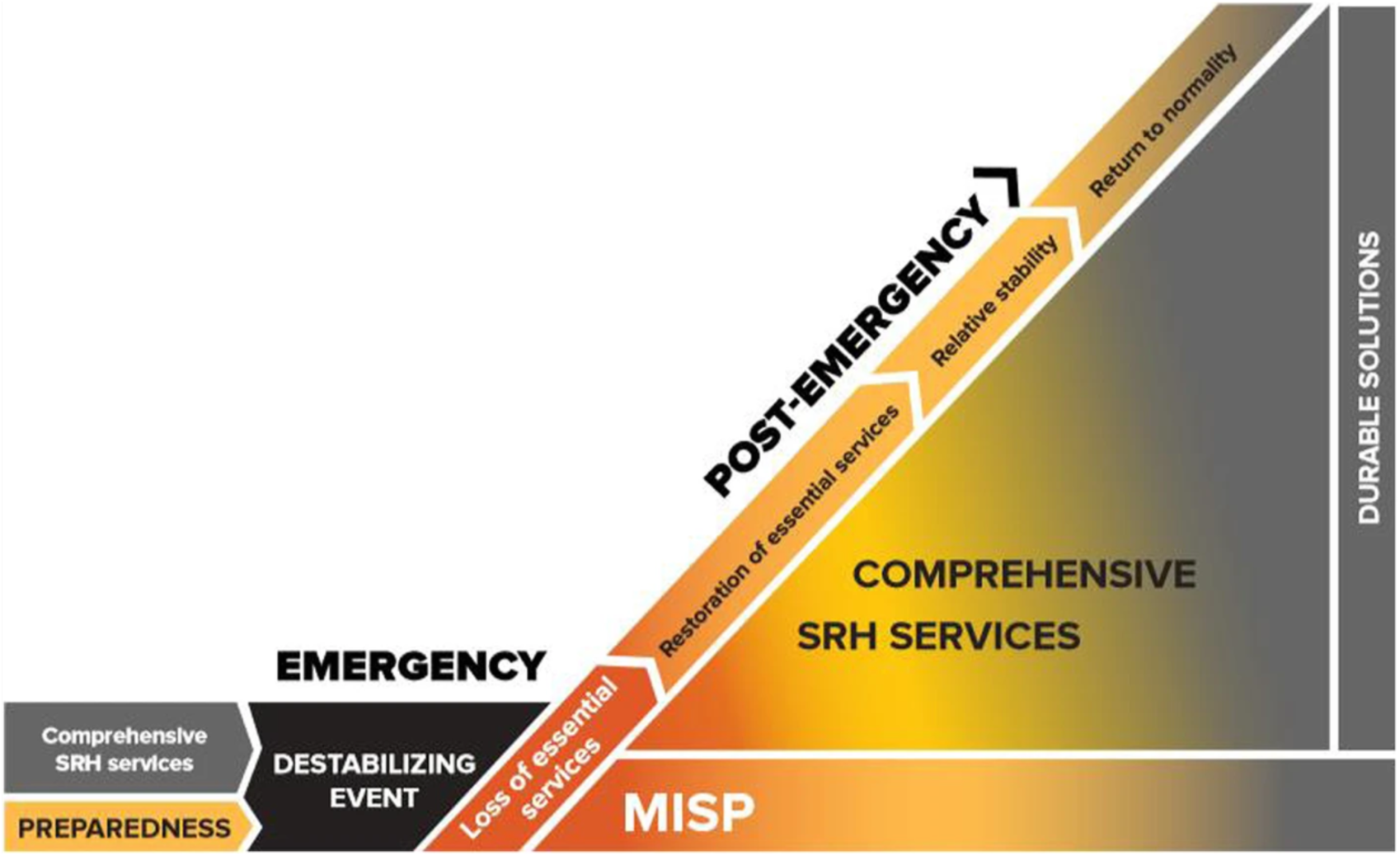
**The Continuum of An Emergency**

“MISP readiness” was also discussed in terms of national, organizational, and individual capacities from the outset of an emergency, such as the Fiji and Tonga context, with a comparison of each country implementing the IPPF’s Sexual and Reproductive Health in Crisis and Post-Crisis Situations (SPRINT) initiative, and flood response in Pakistan through the implementation of the Toolkit for Community Preparedness for SRH and Gender (49,53,91). SRH was more institutionalized in Tonga relative to Fiji, notably in terms of preparedness plans as providers received some trainings on the MISP prior to Cyclone Winston in Fiji, with a more robust “crash course” on the MISP and coordination efforts being delivered after the onset of the cyclone. Conversely, providers in Tonga received trainings prior to Cyclone Gita, encompassing the MISP, long-acting reversible contraceptives (LARCs), and GBV in emergencies, and had stronger, pre-established relationships with the Ministry of Health (MoH). In the aftermath of these emergencies a focus on integrating the MISP into disaster risk management frameworks was discussed, through integration of SRH into national reproductive health policies (53). Although these policies were criticized for not including SRHR, the sections to follow will discuss a separate standalone document that was developed for SRH preparedness, albeit tailored to the COVID-19 pandemic. Preparedness for more comprehensive services in the post-crisis period was also highlighted in the Ready to Save Lives toolkit, specifically for the integration of comprehensive SRH into primary health care services in the immediate aftermath of crisis (91). Similarly, in Indonesia, the case study discussed MISP and disaster preparedness and examined service delivery and how health education disaster and preparedness may be more formalized to incorporate the MISP and maternal and reproductive health in consideration of future crises (63). The authors recommended specific financial targets for the health budget at the national (5%) and regional levels (10%) in order for the government to incorporate preparedness measures into national development plans so that free, quality maternal and reproductive health services may be delivered in the aftermath of crisis (63). Considerations for district, regional, and national structures were also taken into account for flood preparedness efforts in Pakistan, with enhanced collaboration and coordination between communities (via union councils) and the National Disaster Management Agency and the National Health Emergency Preparedness and Response Network (50). While disaster risk infrastructure was already in place in Pakistan, the study highlighted more explicit efforts to institutionalize and integrate SRH-DRR at the sub- national and national levels (50).

### Sub-Theme 1.2. Exit Strategies in Recovery Scenarios

Four publications focused on exit strategies in post-crisis and/or recovery scenarios (52,77–79). In northern Uganda, efforts to dissolve and phase-out the health cluster to sector working groups following a 20-year conflict period were documented as it pertained to SRH services. With the dissolution of the clusters, the authors cited multiple challenges according to those interviewed in the study, including SRH competing with other pressing health demands, the perception that RH consisted only of HIV/AIDS, and the capacity and financial constraints of the government (at national and sub-national levels) to lead and coordinate SRH efforts (52). Save the Children also documented its transition efforts back to the MoH in terms of health facility coverage (financial) in Sudan (encompassing ANC, PNC, and skilled-birth attendance for pregnant women), while in Pakistan, staff hired by the NGO were gradually shifted to government payroll for health programming for ANC, PNC, skilled-birth attendance, FP, and post-abortion care (PAC) (77–79). Challenges in both contexts were noted in terms of capacity (MoH, human resources) and financial sustainability (77,78).

### Theme 2. Transition between minimum and comprehensive services

Building on the concept of the “continuum of an emergency,” the literature documented efforts to transition between minimum and comprehensive SRH services, including feasibility to scale-up or scale-down these services based on context. The scaling down of services was only documented in the literature on COVID-19. Specific measures taken during the COVID-19 pandemic are included in the following section. A total of 13 publications discussed transitions between minimum and comprehensive SRH services (50,53,56,57,63,80,81,83–87,91). Generalized guidance for the transition from minimum to comprehensive services were documented in seven of the publications (80,81,84,85,87,88,91), including the IAWG toolkit with step-by-step instructions to support SRH coordinators to implement objective 6 of the MISP (i.e., “*plan for comprehensive SRH services, integrated into primary health care as soon as possible and work with the Health Sector/Cluster partners to address the six health system building blocks”*) (87). The IAWG toolkit was the first of its kind to guide transition planning up until that point (87). Targeted recommendations for donors were included in funding these transitions, including longer-term, multi-year funding from the emergency period through recovery (85,88).

Five publications provided country examples of MISP implementation during emergency periods where more comprehensive services were interrupted, including Pakistan (50,91), Indonesia (63), Fiji and Tonga (53), and Burkina Faso, Nepal, and South Sudan (56). Conversely, country examples of the transition from minimum to comprehensive services was documented in two different publications, each encompassing the Democratic Republic of Congo (DRC), Yemen, and Bangladesh, with all three countries designated as L3 emergencies during the period under review (57,83). Experiences of implementing the IAWG’s toolkit for transition from MISP to comprehensive SRH included considerations for how to implement this transition, including how to avoid parallel services in populations with high displacement through engagement beyond the MoH, examination of government capacity and structure, and evidence that the acute period of emergency had subsided (57). The WHO evaluation of implementation in DRC, Bangladesh, and Yemen documented key challenges in implementing this transition, including alignment between these efforts and national policies in place (for SRH), the capacity of health workers (and more broadly, implementing organizations), and financial resources available to undertake these efforts and sustain them (83).

### Sub-Theme 2.1. Adaptations during COVID-19, epidemics, and other outbreaks

A total of 12 articles discussed the COVID-19 pandemic and measures taken to adapt or modify services (48,50,51,55,80,82,83,86,87,89,91,92). Of the publications, nine focused on multiple aspects of SRH (50,51,55,80,82,83,87,91,92), two had an explicit focus on contraceptives (48,89) and one on safe abortion care (86). In most fragile contexts, the prioritization of COVID-19 meant the interruption or de-prioritization of SRH services. Given this phenomenon, the literature revealed different measures taken to ensure the delivery of SRH during the pandemic. Beyond COVID-19 implications, the literature also highlighted broader applications to future pandemics, epidemics, and infectious disease outbreaks. Approximately half of the publications were generalized across fragile contexts, with six providing global operational guidance (80,81,86,87,91,92) and one peer-reviewed publication providing examples of the impact of COVID-19 on contraceptive use across the HDN (48). Additionally, one publication provided guidance specific to the South and Southeast Asia and Pacific regions on the integration of SRH in COVID- 19 national pandemic preparedness plans and recommended country-level adaptations across the region to ensure SRH service delivery during the pandemic (82). Country-specific examples of COVID-19 and its impact on service delivery encompassed Pakistan (50,91); Nigeria (51,55); Afghanistan, Colombia, Iraq, Somalia, South Sudan, Syria, Venezuela, Zimbabwe (55); DRC, Yemen, and Cox’s Bazar, Bangladesh (55,83).

Drawing from the same “continuum of an emergency” concept, SRH service continuity amidst the COVID-19 pandemic was a prominent theme in the literature. This included the IAWG’s toolkit and facilitation guide (for SRH coordinators) on the transition from minimum to comprehensive services alongside essential emergency services and infection prevention and control (IPC) (87) and its supplemental guide on adolescent SRH outlining barriers to access (disability, stigma, cultural norms) and recommendations for increasing SRH uptake, such as implementing national training curricula for SRH in countries where the MoH has not already done so (90). The International Rescue Committee and IAWG created a checklist for maintaining access to safe abortion care citing Zika virus, Ebola, and COVID-19 as examples (86), while the Ready to Save Lives Preparedness Toolkit focused on SRH policy integration (as discussed in the previous section), streamlining the availability of comprehensive SRH services within 3-6 months of an outbreak, and enhancing coordination between SRH and outbreak sectors through country-level preparedness plans (91). Similarly, guidance from the WHO, UNFPA, and UNICEF emphasized the service continuity through country-specific adaptations, underscoring the need to maintain existing national standards of care, rather than eliminating these services and contributing to backslides in health (82).

Adaptive measures highlighted in publications included the redesign, scaling down, and decentralization of health services and stocks (51,55,82,89,91). Other notable strategies included a shift in service modalities, such as task-shifting, the reliance on community-based service delivery, and telemedicine (48,50,51,80,82,83,89,91,92), and self-care interventions, including the provision of LARCs and mid-upper arm circumference (MUAC) measurement trainings (in cases of malnutrition) (51). Engagement with private sector pharmacies and health facilities to provide FP services was also reported as a strategy given the burden of COVID-19 on public health facilities (48,89). The inability to maintain FP service delivery during the pandemic was highlighted as a challenge for multiple reasons, including a lower demand for services, the repurposing or closure of health facilities, supply chain interruptions, and the de-prioritization of FP services citing the perception of FP interventions as non-lifesaving relative to other health services (48,89). In Nigeria, service providers reported some MNCH activities and/or entire programs were eliminated due to the delay in receiving COVID-19 funding and the need to re-channel MNCH funds to cover these services (55). Lastly, other secondary health outcomes were reported across multiple settings by Rodo et al. because of the pandemic, including improvements in hygiene and a reduction in disease (beyond COVID-19 infections) due to enhanced IPC and WASH (Water, Sanitation & Hygiene) measures (55). Conversely, other consequences defined as “negative secondary effects” were reported, such as the impact on SRH and GBV services due to the elimination/reduction of community outreach services, in addition to equity concerns for patients who did not have access to technology, who in turn, were unable to utilize telemedicine (55).

### Theme 3. Health Systems Strengthening

Health Systems Strengthening (HSS) has traditionally been viewed as a development activity, often deemed less feasible in humanitarian settings. The literature documented examples of HSS in acute and protracted crisis, along with guidance on HSS measures that may be undertaken prior to a post-conflict, recovery scenario. A total of 12 publications focused on HSS (50,57,60,64,68,71,74,75,77–79,91). These included more generalized global guidance (91), along with publications encompassing South Sudan (60,64), Ethiopia (71), Somalia (74), DRC (57,74,75), Pakistan and Sudan (50,77–79,91), and Yemen and Cox’s Bazar, Bangladesh (57).

Different publications emphasized working closely with the MoH, including MoH-supported personnel and facilities to strengthen the health system in protracted settings. These included the provision of FP services in MoH-supported health facilities in North and South Kivu, DRC (75); scale up of FP services in select government health facilities in Pakistan (78,79); transitioning health facilities and activities to the MoH in Sudan (from an international NGO) (77,79); and expanding health information systems of the MoH to include disaggregated data for contraceptive and PAC services in DRC and Somalia (74). Beyond engagement with the MoH, Tanabe et al. emphasized engagement with other national and sub-national agencies in SRH resilience-building efforts in Pakistan (as detailed above) (50), while Tran and colleagues stressed the importance of including other critical ministries, such as finance (for health financing), home affairs (for programming involving displaced populations), and education (for training the health workforce and institutionalizing SRH services) as documented in Yemen, DRC, and Bangladesh (57). Feasibility of HSS, was examined in context of South Sudan, where Valadez et al. emphasized a non-uniform, decentralized approach to HSS across different states within the country, given that fragility varied from state to state (60). Odhiambo et al. looked at the health system’s resilience to shocks and stresses in as it pertained to MNCH services through a measurable resilience index with stress indicators. It was found that states with high stress actually had higher resilience measures due to the presence of humanitarian and development actors (along with security forces), and in turn, more effective governance (64).

### Sub-theme 3.1. Health Systems Strengthening through Workforce Development

A total of 18 publications focused on task-shifting and capacity strengthening (48–50,53,55,56,60,62,66,70,74,77,78,82,83,89,91,92). This included generalized examples across different contexts (48,49,55,66,82,89,91,92), and those from the DRC (74,83), Pakistan (50,78), Sudan (77), Somalia (74), South Sudan (56,60,62), Fiji and Tonga (53), Burkina Faso and Nepal (56), Burundi and northern Uganda (70), and Bangladesh and Yemen (83). The functioning of any health system is contingent upon a capable, local workforce. Investment in this workforce is a critical component of HSS, and this is imperative in fragile contexts where health cadre must be equipped with the knowledge and resources to respond to shocks and stresses. Significant challenges were highlighted in the literature in ensuring adequate human resources for health in fragile settings, including weakened health infrastructure and limited government investments in the national health budget and medical education system. Within the literature, task-shifting and capacity strengthening were identified as key measures to addressing the challenges of education, training, recruitment, and retention of health cadre in fragile settings. As discussed previously, COVID-19 necessitated task-shifting of SRH services to lower level health cadre given the prioritization of essential health services, such as shifting contraceptive provision to community health workers (48,89) and utilizing traditional birth attendants (TBAs) (in contexts like Nigeria and Bangladesh) in place of facility-based deliveries by skilled birth attendants (SBAs, e.g., nurses, midwives) given women were often unable to access health facilities during the pandemic (55). These approaches aligned with broader guidance materials on task-shifting and capacity strengthening which were developed (or underway) prior to the pandemic (84,90,92) and those which emerged during COVID-19 (80,82,83,89).

Country-specific examples of capacity strengthening efforts were documented in the literature, including a clinical refresher training program for the clinical management of sexual violence and manual vacuum aspiration implemented in Burkina Faso, Nepal, and South Sudan, which authors noted that could not replace more comprehensive capacity strengthening strategies and plans led by each respective government (56). In Fiji and Tonga, capacity strengthening took place at the individual and institutional levels on SRH in emergencies, but also in the “role flexibility” of health cadre who had to deliver services outside of their duties and scope of SRH (53). In DRC and Somalia, trainings were provided on PAC and FP to allow midlevel providers to carry out certain services and allowing these services to be offered at lower-level healthcare facilities (74). In South Sudan, task-shifting was cited as a short-term intervention to address the dire shortage of human resources for health amidst protracted conflict, though key challenges to long-term feasibility were cited, including discrepant (and inconsistent) salaries for government health staff and the reliance on NGOs for service delivery (60,62).

Barriers to retention were identified, including insufficient supplies, gaps in the referral pathway, and limited supervision and continuing education (66). Miyake et al. looked at the role of midwives and how SBAs were recruited and retained through different “retention enablers,” such as community-based deployment to ensure sustainable presence of midwives in their own communities (66). In Burundi and northern Uganda, task-shifting of obstetric care from SBAs to TBAS was used as an adaptive measure given the shortage of human resources during the period of conflict in each setting, with the authors citing that TBAs were often the first point of entry for health in communities. In the post-conflict period, TBAs in both settings were initially not permitted to perform such activities by their respective governments, though Burundi later re-assigned TBAs the role of “birth companions,” particularly in rural areas. In northern Uganda, TBAs have not been included or re-integrated in the healthcare system, though it was noted that many TBAs still provided such services informally (70). Other adaptations have been made in post-conflict periods, as documented by Chol et al. in terms of contracting foreign providers (from Cuba and China) and providing midwifery and training courses (Eritrea), in utilizing health extension workers (Ethiopia), and in increasing the number of community health workers (Mozambique, Rwanda) (69).

### Sub-theme 3.2. Other Health System Strengthening Reforms

From the literature, 19 publications encompassed financing, coordination, and leadership and governance (48,50,52,54,57–59,69,71,81,82,84,85,87–92). Other reforms were discussed in the literature, drawing from the health system building blocks. Reforms in terms of health system components were often discussed in tandem, specifically for governance (policies) and financing. Broader financing recommendations to donors (especially in protracted settings) were made in order to address shorter funding cycles (62), increase development funding for SRH beyond HIV/STI in settings where there is concurrent humanitarian funding (54,58), and strengthen the transition from the MISP to more comprehensive SRH services through continuous, flexible, and multi-year funding (as discussed in Theme 2) (85,88). Other recommendations included rapid funding for emergencies at their onset (85), including for the MISP (88).

In Ethiopia, efforts to attain and expand UHC through the integration of SRHR in national policies and the UHC package were documented through detailed costing plans consisting of domestic financing (by government), out-of-pocket expenditures, and external development assistance (71). Berhan et al. noted that although the UHC package listed various SRHR services greater progress was seen in MCH and FP services due to political and financial commitments given their explicit linkage to the Millenium Development Goals. Conversely, other services were either not included in the UHC package or not fully implemented (e.g., treatment for FGM complications, comprehensive sexuality education, and sexual health services, including treatment for infertility and reproductive cancers) due to pervasive barriers (financial, gender, and sociocultural norms) (71). In Chol et al., the authors outlined various reforms to address maternal mortality, which encompassed health financing and decentralization of health governance structures in Angola, Eritrea, Ethiopia, Mozambique, and Rwanda following conflict during the period of 1990-2015 (in addition to human resource reforms outlined previously) (69). In the five countries reviewed, health financing reforms outlined the ways in which countries shifted from external aid dependence to some form of cost-recovery post-conflict, including fee retention policies for health facilities (Ethiopia), performance-based financing (Rwanda), community-based health insurance schemes (Rwanda, Eritrea, Ethiopia), pooled donor financing (Eritrea, Mozambique), and an increase in out-of-pocket expenditures and government coverage (Angola) (69).

As discussed in earlier themes, coordination efforts were documented in Pakistan (50), northern Uganda (52), and DRC, Bangladesh, and Yemen (57), along with broader guidance on coordination across different contexts (81,82,84,87,90–92). Coordination and collaboration with the private sector were also documented in the literature, particularly in contexts with a fragile health system and weakened public sector with significant gaps in health services (including those delivered by traditional humanitarian actors). Publications that focused on contraceptives outlined the use of vouchers for LARCs and permanent methods in Yemen and Pakistan (59), engagement with private pharmacies and health facilities to distribute LARCs to clinicians at these sites who could insert/remove them given the burden on public health facilities during COVID-19 and supply chain interruptions across multiple fragile contexts (48,89). Boddam-Whetham and colleagues documented public-private sector coordination and engagement, and cited financial barriers to accessing LARCs in Yemen, as only the wealthiest segment of the population (residing in the larger cities) was able to access these methods (15.9%) relative to the poorest (1.6%) (59). By contracting out to the private sector, the program was considered a precursor to private health insurance since it entailed some form of cost recovery for the health system (59).

## DISCUSSION

Across the 45 publications included within this scoping review, 39 LMIC settings were included, representing just over 40% of all LMICs that were classified as fragile at some point during the period between 1990-2023 (33–36). In line with the continuing evolution of nexus terminology and the nascency of its operationalization for the health sector, the literature showcased a diverse set of concepts and themes on the nexus, though this terminology was not always explicitly used.

While the aim of this scoping review was to synthesize literature related to the HDpN and SRMNCAH interventions in fragile settings, the bulk of the relevant literature focused on the HDN and SRH. No included studies focused on peace aspects and very few focused on child and adolescent health. The gaps in the literature on the peace component of the HDpN may have been due to the nature of peace interventions, which have often been interpreted to be entrenched in the political realm, and therefore are not published in the health databases we searched. Peace programming has also been presumed to challenge the “identity” of stakeholders working across the nexus (e.g., humanitarian, development), and perceived by some to be at odds with the humanitarian principles, namely, those of neutrality, impartiality, and independence (93–95). In terms of child health, the limited focus on these interventions within the HDN may be due to the existing buy-in on the importance of these interventions across the emergency continuum, as these have been explicitly outlined as priorities in humanitarian interventions, the SDGs, and targets for achieving UHC (96,97). Conversely, adolescent health has only more recently been emphasized in terms of specific, targeted guidance (90,98), though global evidence points to a great need for adolescent-tailored services (99), including across the HDN.

The expansion of SRH preparedness measures in fragile settings remains essential given the current trajectory of climate emergencies, which are estimated to contribute to 14.5 million deaths and 1.2 billion displacements by 2050 (17–19). Country examples of the integration of SRH-DRR in this review provide contextualized recommendations for how to feasibly incorporate them within different countries and regions according to varied and changing contexts, including explicit financial targets for integrating preparedness measures into development plans, as we saw in the Sajow et al. study in Indonesia (63). These examples build on the current, generalized—and somewhat generic—guidance materials on SRH-DRR integration (100,101). In this regard, the literature addressed a current gap, as the overarching global framework for DRR (i.e., the Sendai Framework) does not include targeted recommendations by sector (including health and SRH). Therefore, it should be useful for practitioners to have real-world implementation efforts charted in this review (20). More regional approaches (without SRH included) have been developed given multiple countries may be collectively pre-disposed to certain shocks, such as the Sahel (e.g., drought, food insecurity) and Pacific regions (e.g., cyclones, earthquakes) (102,103). While regional adaptations to the Sendai Framework exist, DRR that formally incorporates SRH remains essential. The intersection of SRH and climate shocks in at-risk contexts, as presented in the findings, also supplements the limited literature base on SRH and anticipatory action for climate shocks in fragile contexts. This includes the 2023 global review of SRHR in national climate commitments undertaken by UNFPA that examined 119 different countries and highlighted the limited progress in SRHR integration along with gaps in addressing gender, GBV, and harmful practices that are all fundamental to SRHR (104).

In terms of SRH implementation across the nexus, the ability to flexibly transition between minimum and comprehensive services was an important feature highlighted in the literature. Beyond this, it is clear that adaptive measures are necessary for service continuity during crisis and. Both of these notions build on previous efforts to safeguard SRH services, regardless of context (105,106). The country examples presented in this scoping review have further contextualized the existing guidance (e.g., the MISP, IAWG toolkit) to transition between services across a diverse array of contexts, including efforts to expand minimum services in L3/active scale up emergencies (57,83). These implementation examples are a useful resource given that they address contextual nuances, which may translate to other settings. There is still a need to document flexible SRH transitions in complex operational environments, and create concrete guidance to navigate these challenges, including pervasive issues like resistant sociocultural and religious norms, violence toward health care workers, sanctions, and the presence of state (MoH) and de facto (non-state) health authorities (36,40,107,108). These factors intersect with health governance and coordination, and greatly influence the feasibility of implementation in such restrictive operational environments.

The COVID-19 pandemic proved to be a significant disruptor of SRH services, and a key inflection point in collaboration between humanitarian and development practitioners. The demonstrated use of task-shifting, decentralization, and telemedicine are practical tools that may be adopted to maintain service delivery, though telemedicine may not always be feasible in settings where there is limited access to technology (55). To date, there are limited examples in the literature on scaling down service delivery while still preserving access to essential SRH care. Though GBV was not the focus of this review, the elimination of SRH community outreach services during the COVID-19 pandemic interrupted access to these along with referral pathways for GBV services (55). With the health sector recognized as a critical entry point for addressing GBV, these measures may have contributed to what UN Women has termed, the “shadow pandemic,” which entailed a surge in the rates of violence toward women and girls during the COVID-19 pandemic (109).

As a prominent theme in this review, health systems strengthening was identified as an essential measure across the emergency continuum. With more crises extending for years, the wider literature has emphasized the necessity of health systems strengthening for reaching the SDGs, even in fragile settings (23). Similarly, there is a growing interest in the resilience of health systems, including recommendations on how to enhance this resilience in fragile settings (110,111). The substantial focus on workforce development in the findings underscores the criticality of human resources in the health system, particularly the local workforce. The emphasis on task-shifting to lower-level health cadre and capacity strengthening complements the broader literature on adaptations for service delivery in LMICs in the absence or scarcity of qualified health professionals (e.g., physicians, nurses, midwives). By 2030, the WHO has projected that there will be a health workforce shortage of 14 million, mostly in countries in Africa, the Middle East, and Asia and the Pacific (35,112). In these (human) resource-deprived settings, the use of community-level health cadre (e.g., TBAs, extension health workers, community outreach) is critical and builds on two essential premises, as evidenced by the broader literature. First, community health workers are a critical entry point for communities seeking health care, including, and especially at the onset of an emergency (i.e., frontline or first responders) (113–115). Second, community health is an important component of primary health care within the health system, and in turn, primary health care is the “predominant pathway for achieving UHC” and a “prerequisite for achieving the Sustainable Development Goals” as Perry and Sachs have emphasized in their call for greater investment in primary health care in LMIC settings (97).

Other bottlenecks highlighted in this review include the renumeration of health cadre and institutionalization of lower-level health workers, especially at the community level. From a policy standpoint, there should be formal measures to re-integrate and/or institutionalize community health roles within national health policies, and subsequently, the health system, regardless of the level of fragility. These efforts are necessary for the expansion or scale up of human resource capacity, including in recovery scenarios, and require detailed costing plans that align with health policies in place. Furthermore, health plans should be designed in tandem with cost projections, in order to make sure they are feasible to implement. Collins et al. have documented efforts to calculate the costs of integrating community health packages and examined key aspects of costing for community health packages, including the types of services to be included (e.g., FP, ANC, PNC, HIV), and whether or not community health cadre are accredited and renumerated (116). Similar investment cases for community health have been made at the country level, such as South Sudan’s Boma Health Worker initiative (117) and global recommendations using Liberia, Ethiopia, and Pakistan as examples (118). In terms of midwives and other SBAs, “retention enablers,” such as community-linked deployment contributes to a more sustainable workforce are needed (66). These measures align with the global efforts to invest in SBAs, in terms of education, supervision, and the regulatory (legal) framework for SBAs within the health system (66,119). Just as with community health workers, midwives (and SBAs at large), have been deemed essential for achieving UHC and the SDGs (119).

As it stands, more sustainable health financing in fragile, LMICs remains a key priority given the limited resources (human, financial), modest government investments in national health budgets, and significant reliance on external sources of funding to subsidize health care costs. Without adequate cost-recovery mechanisms in place for health services (and health providers) in these settings, sustainability is difficult and there will continue to be a reliance on humanitarian assistance in the short-term, while in the longer-term, development assistance will be limited in its effectiveness. In addition to these considerations, the literature has underscored the need for more innovative financing approaches for humanitarian assistance given current funding cycles are 6-12 months on average relative to 3-5+ years for development programming. The discrepancy in funding cycles for humanitarian and development interventions has made alignment and complementarity difficult. Though there are more examples of multi-year emergency awards (with a broader resilience focus) emerging (e.g., USAID, European Commission), these approaches are still somewhat in their infancy and have not yet been mainstreamed among institutional donors (120,121). There are examples of a shift from reliance on external assistance (both human and financial resources) through the piloting/implementation of different cost-recovery models (e.g., performance- or results-based financing), engagement with the private sector (e.g., vouchers), and the development of more deliberate and well-conceived exit strategies in the literature (59,69,77,78). However, the proliferation of more complex, protracted crises will likely continue to have a significant impact on the economic conditions and financial viability in many of these fragile settings.

### Strengths and Limitations

This review summarizes global evidence on SRH and the HDN and provides concrete guidance on how to operationalize this for SRH interventions. It should serve as a valuable contribution to global evidence. In addition, this review attempted to move past HDN jargon and document what the nexus is no matter how it is described, including successes and challenges in implementation. This review provides a foundation for SRH practitioners and researchers looking to examine the operationalization of the HDN for SRH in fragile settings given a scoping review on this topic does not yet exist in the literature.

This review had some limitations. Given the continuing evolution of the nexus terminology and its more nascent application to the health sector, including SRH, there may have been studies that were not captured through the search terms used in this review. In terms of grey literature databases, these were skewed toward humanitarian interventions, except a few that aggregated resources from humanitarian and development practitioners. This may be due to more documentation by humanitarian practitioners, or at least more efforts to compile these resources in one place. Given that this study did not account for GBV, there may have been other studies that could have been included, as SRHR and GBV are inextricably linked. There remains a wide interpretation of the nexus approach given the terminology differed between publications and sources, therefore, the studies included do not represent all such efforts to operationalize the nexus for SRH. Finally, this review only included studies published in English, therefore, relevant studies in other languages (French, Arabic, Spanish) may have been excluded that could have added to the body of literature on the HDN and SRH interventions.

## CONCLUSION

This scoping review demonstrates a great need and appetite to operationalize the HDN for SRH interventions in fragile settings. The integration of SRH within DRR and national health policies, especially for countries (and regions) predisposed to recurrent climate shocks is necessary. Country examples presented on the transition between minimum and comprehensive SRH services supplement the existing guidance in the literature and offer further contextualization. Adaptive measures taken during the COVID- 19 pandemic, including the decentralization of health services, shift to community-based service modalities, and telemedicine may inform future pandemic and outbreak response. HSS and resilience-building efforts remain necessary in fragile settings and add to the growing body of evidence on these topics. The use of task-shifting to lower-level health cadre (and capacity strengthening) offers solutions for resource-deprived, LMIC settings where significant health workforce shortages are projected to increase by 2030. Formal measures to institutionalize and integrate these cadre within the health system will contribute to more adequate primary health care in the short-term, and in turn, help achieve UHC and the SDGs, in the medium- and long-term. Health financing remains an essential component of operationalizing the HDN, including multi-year funding considerations, engagement with the private sector, feasible cost-recovery mechanisms, and more deliberate, well-conceived exit strategies. Together, these may help shift reliance on external assistance (both humanitarian and development). Lastly, there should be more alignment between health planning and costing projections to implement these plans to ensure sustainability.

## Data Availability

All data produced in the present work are contained in the manuscript.

## ACKNOWLEDGEMENTS

We sincerely thank Shannon Doocy and Shannon Frattaroli from the Johns Hopkins Bloomberg School of Public Health for their guidance throughout this research and for their extensive review of this manuscript. We thank Donna Hesson for her guidance and support during the development of the search strategy for this scoping review. Amany Qaddour would also like to thank the Center for Human Rights and Humanitarian Studies, Watson Institute for International and Public Affairs at Brown University for generously providing a research stipend for student research assistants Stefany Lazieh and Ava Ward during her time there as a visiting scholar in 2022-2023.

## SUPPORTING INFORMATION

▪ S1 File. Preferred Reporting Items for Systematic Reviews and Meta-Analyses Extension for Scoping Reviews (PRISMA-ScR) Checklist
▪ S2 File. Peer-Reviewed Database Search Terms
▪ S3 File. Grey Literature Sources
▪ S4 File. Low- and Middle-Income Countries by Fragility Inclusion Criteria

