## Supplementary material for "The Humanitarian-Development Nexus and Sexual and Reproductive Health Interventions in Fragile Settings: A Scoping Review": S2 File.docx

**S2 File. Peer-Reviewed Database Search Terms**

**CINAHL Database Search**

**Search Terms**

- Search Term/Concept 1—LMICs, fragile and conflict-affected states
- Search Term/Concept 2—humanitarian development peace nexus (HDPN)
- Search Term/Concept 3—maternal and child health/sexual and reproductive health (SRMNCAH)

((MH “afghanistan”  OR MH “ angola”  OR MH “ armenia”  OR MH “ azerbaijan”  OR MH “ bangladesh”  OR MH “ benin”  OR MH “ bolivia”  OR MH “ bosnia and herzegovina”  OR MH “ burkina faso”  OR MH “ burundi”  OR MH “ cambodia”  OR MH “ cameroon”  OR MH “ central african republic”  OR MH “ chad”  OR MH “ Chechnya”  OR MH “ chile”  OR MH “ colombia”  OR MH “ comoros”  OR MH “ democratic republic of the congo”  OR MH “ congo”  OR MH “ cote d’Ivoire”  OR MH “ cuba”  OR MH “ djibouti”  OR MH “ east timor”  OR MH “ ecuador”  OR MH “ el salvador”  OR MH “ eritrea”  OR MH “ eswatini”  OR MH “ ethiopia”  OR MH “ fiji”  OR MH “ gambia”  OR MH “ grenada”  OR MH “ guatemala”  OR MH “ guinea”  OR MH “ guinea-bissau”  OR MH “ guyana”  OR MH “ haiti”  OR MH “ honduras”  OR MH “ indonesia”  OR MH “ iran”  OR MH “ iraq”  OR MH “ jordan”  OR MH “ kenya”  OR MH “ Kiribati”  OR MH “ democratic people’s republic of korea”  OR MH “ republic of korea”  OR MH “ kosovo”  OR MH “ kyrgyzstan”  OR MH “ laos”  OR MH “ lebanon”  OR MH “ lesotho”  OR MH “ liberia”  OR MH “ libya”  OR MH “ republic of north macedonia”  OR MH “ madagascar”  OR MH “ malawi”  OR MH “ mali”  OR MH “ micronesia”  OR MH “ mauritania”  OR MH “ mozambique”  OR MH “ myanmar”  OR MH “ namibia”  OR MH “ nepal”  OR MH “ netherlands antilles”  OR MH “ nicaragua”  OR MH “ niger”  OR MH “ nigeria”  OR MH “ occupied Palestinian territory”  OR MH “ pakistan”  OR MH “ papua new guinea”  OR MH “ peru”  OR MH “ philippines”  OR MH “ puerto rico”  OR MH “ russia”  OR MH “ sao tome and principe”  OR MH “ senegal”  OR MH “ sierra leone”  OR MH “ Solomon islands”  OR MH “ somalia”  OR MH “ south sudan”  OR MH “ sri lanka”  OR MH “ sudan”  OR MH “ syria”  OR MH “ tajikistan”  OR MH “ tanzania”  OR MH “ timor-leste”  OR MH “ togo”  OR MH “ tonga”  OR MH “ turkey”  OR MH “ Tuvalu”  OR MH “ uganda”  OR MH “ ukraine”  OR MH “ uzbekistan”  OR MH “ vanuatu”  OR MH “ venezuela”  OR MH “ west bank and gaza”  OR MH “ west timor”  OR MH “ yemen”  OR MH “ zambia”  OR MH “ zimbabwe” ) OR (AB “afghanistan”  OR AB “ angola”  OR AB “ armenia”  OR AB “ azerbaijan”  OR AB “ bangladesh”  OR AB “ benin”  OR AB “ bolivia”  OR AB “ bosnia and herzegovina”  OR AB “ burkina faso”  OR AB “ burundi”  OR AB “ cambodia”  OR AB “ cameroon”  OR AB “ central african republic”  OR AB “ chad”  OR AB “ Chechnya”  OR AB “ chile”  OR AB “ colombia”  OR AB “ comoros”  OR AB “ democratic republic of the congo”  OR AB “ congo”  OR AB “ cote d’Ivoire”  OR AB “ cuba”  OR AB “ djibouti”  OR AB “ east timor”  OR AB “ ecuador”  OR AB “ el salvador”  OR AB “ eritrea”  OR AB “ eswatini”  OR AB “ ethiopia”  OR AB “ fiji”  OR AB “ gambia”  OR AB “ grenada”  OR AB “ guatemala”  OR AB “ guinea”  OR AB “ guinea-bissau”  OR AB “ guyana”  OR AB “ haiti”  OR AB “ honduras”  OR AB “ indonesia”  OR AB “ iran”  OR AB “ iraq”  OR AB “ jordan”  OR AB “ kenya”  OR AB “ Kiribati”  OR AB “ democratic people’s republic of korea”  OR AB “ republic of korea”  OR AB “ kosovo”  OR AB “ kyrgyzstan”  OR AB “ laos”  OR AB “ lebanon”  OR AB “ lesotho”  OR AB “ liberia”  OR AB “ libya”  OR AB “ republic of north macedonia”  OR AB “ madagascar”  OR AB “ malawi”  OR AB “ mali”  OR AB “ micronesia”  OR AB “ mauritania”  OR AB “ mozambique”  OR AB “ myanmar”  OR AB “ namibia”  OR AB “ nepal”  OR AB “ netherlands antilles”  OR AB “ nicaragua”  OR AB “ niger”  OR AB “ nigeria”  OR AB “ occupied Palestinian territory”  OR AB “ pakistan”  OR AB “ papua new guinea”  OR AB “ peru”  OR AB “ philippines”  OR AB “ puerto rico”  OR AB “ russia”  OR AB “ sao tome and principe”  OR AB “ senegal”  OR AB “ sierra leone”  OR AB “ Solomon islands”  OR AB “ somalia”  OR AB “ south sudan”  OR AB “ sri lanka”  OR AB “ sudan”  OR AB “ syria”  OR AB “ tajikistan”  OR AB “ tanzania”  OR AB “ timor-leste”  OR AB “ togo”  OR AB “ tonga”  OR AB “ turkey”  OR AB “ Tuvalu”  OR AB “ uganda”  OR AB “ ukraine”  OR AB “ uzbekistan”  OR AB “ vanuatu”  OR AB “ venezuela”  OR AB “ west bank and gaza”  OR AB “ west timor”  OR AB “ yemen”  OR AB “ zambia”  OR AB “ zimbabwe”))

**AND: Humanitarian-Development-Peace Nexus Concepts (Search Term 2)**

((MH “humanitarian development” OR MH “ humanitarian development peace” OR MH “ humanitarian development peace nexus “ OR MH “ double nexus” OR MH “ triple nexus” OR MH “ disaster relief “ OR MH “ disaster risk reduction “ OR MH “ disaster-affected” OR MH “ humanitarian development divide OR MH “ humanitarian development continuum” OR MH “ humanitarian development gap” OR MH “ relief to development “ OR MH “ linking relief, rehabilitation and development” OR MH “ fragile state” OR MH “ fragility” OR MH “ fragile setting” OR MH “ fragile country” OR MH “ conflict-affected” OR MH “ war-torn” OR MH “ war-affected” OR MH “ displacement setting” OR MH “ refugee setting” OR MH “ emergency relief “ OR MH “ complex emergency “ OR MH “ protracted crises”) OR (AB “humanitarian development” OR AB “ humanitarian development peace” OR AB “ humanitarian development peace nexus “ OR AB “ double nexus” OR AB “ triple nexus” OR AB “ disaster relief “ OR AB “ disaster risk reduction “ OR AB “ disaster-affected” OR AB “ humanitarian development divide OR AB “ humanitarian development continuum” OR AB “ humanitarian development gap” OR AB “ relief to development “ OR AB “ linking relief, rehabilitation and development” OR AB “ fragile state” OR AB “ fragility” OR AB “ fragile setting” OR AB “ fragile country” OR AB “ conflict-affected” OR AB “ war-torn” OR AB “ war-affected” OR AB “ displacement setting” OR AB “ refugee setting” OR AB “ emergency relief “ OR AB “ complex emergency “ OR AB “ protracted crises”))

**AND: SRMNCAH Interventions (Search Term 3)**

MH “maternal-child health” OR MH “maternal-child welfare” OR MH “child health” OR MH “child health services” OR MH “adolescent health” OR MH “adolescent health services” OR MH “maternal health services” OR MH “maternal health services” OR MH “immunization” OR MH “vaccines” OR MH “sexual health” OR MH “reproductive health” OR MH “right to health” OR MH “reproductive rights” OR MH “delivery, obstetric” OR MH “obstetric service” OR MH “obstetric care” OR MH “surgery, obstetrical” OR MH “obstetric patients” OR MH “gynecology” OR MH “gynecologic examination” OR MH “surgery, gynecologic” OR MH “gynecologic care” OR MH “contraception” OR MH “contraceptives, postcoital” OR MH “hormonal contraception” OR MH “contraceptives, oral combined” OR MH “contraceptive agents, male” OR MH “diaphragms, contraceptive” OR MH “family planning” OR MH “family planning, natural” OR MH “birth intervals” OR MH “intrapartum care” OR MH “prenatal care” OR MH  “perinatal care” OR MH “postnatal care”

OR “maternal-child health” OR AB “maternal-child welfare” OR AB “child health” OR AB “child health services” OR AB “adolescent health” OR AB “adolescent health services” OR AB “maternal health services” OR AB “maternal health services” OR AB “maternal child health” OR AB “maternal and children health” OR AB “newborn health” OR AB “neonatal health”AB “immunization*” OR AB “vaccines” OR AB “sexual health” OR AB “reproductive health” OR AB “right to health” OR AB “reproductive right*” OR AB “sexual and reproductive health” OR AB “sexual and reproductive health and right*” OR “gynecological” AB “obstetric” OR AB “obstetric service” OR AB “obstetric care” OR AB “obstetrical” OR AB “obstetric patient*” OR AB “gynecolog*” OR AB “gynecologic examination” OR AB “gynecologic surgery” OR AB “gynecologic care” OR AB “contraception” OR AB “contraceptives, postcoital” OR AB “hormonal contraception” OR AB “contraceptives, oral combined” OR AB “contraceptive agents, male” OR AB “diaphragms, contraceptive” OR AB “family planning” OR AB “family planning, natural” OR AB “birth spacing” OR AB “birth spacing” OR AB “healthy timing and spacing of pregnanc*” OR AB “intrapartum care” OR AB “prenatal care” OR AB  “perinatal care” OR AB “postnatal care”

**SEARCH RESULTS: 432,020**

|  | Search results |
| --- | --- |
| Search term 1—mh | 206, 083 |
| Search term 1—abstract |  |
| Search term 2—mh | 4,216 |
| Search term 2—abstract |  |
| Search term 3—mh | 170,027 results |
| Search term 3—abstract | 86,091 results |
| 1 AND 2 AND 3  (mh and tw) | **64** |

**PubMed Database Search**

(((afghanistan[MeSH] OR angola[MeSH] OR armenia[MeSH] OR azerbaijan[MeSH] OR bangladesh[MeSH] OR benin[MeSH] OR bolivia[MeSH] OR bosnia and herzegovina[MeSH] OR burkina faso[MeSH] OR burundi[MeSH] OR cambodia[MeSH] OR cameroon[MeSH] OR central african republic[MeSH] OR chad[MeSH] OR Chechnya[MeSH] OR chile[MeSH] OR colombia[MeSH] OR comoros[MeSH] OR democratic republic of the congo[MeSH] OR congo[MeSH] OR cote d’Ivoire[MeSH] OR cuba[MeSH] OR djibouti[MeSH] OR east timor[MeSH] OR ecuador[MeSH] OR el salvador[MeSH] OR eritrea[MeSH] OR eswatini[MeSH] OR ethiopia[MeSH] OR fiji[MeSH] OR gambia[MeSH] OR grenada[MeSH] OR guatemala[MeSH] OR guinea[MeSH] OR guinea-bissau[MeSH] OR guyana[MeSH] OR haiti[MeSH] OR honduras[MeSH] OR indonesia[MeSH] OR iran[MeSH] OR iraq[MeSH] OR jordan[MeSH] OR kenya[MeSH] OR Kiribati[MeSH] OR democratic people’s republic of korea[MeSH] OR republic of korea[MeSH] OR kosovo[MeSH] OR kyrgyzstan[MeSH] OR laos[MeSH] OR lebanon[MeSH] OR lesotho[MeSH] OR liberia[MeSH] OR libya[MeSH] OR republic of north macedonia[MeSH] OR madagascar[MeSH] OR malawi[MeSH] OR mali[MeSH] OR micronesia[MeSH] OR mauritania[MeSH] OR mozambique[MeSH] OR myanmar[MeSH] OR namibia[MeSH] OR nepal[MeSH] OR netherlands antilles[MeSH] OR nicaragua[MeSH] OR niger[MeSH] OR nigeria[MeSH] OR occupied Palestinian territory[MeSH] OR pakistan[MeSH] OR papua new guinea[MeSH] OR peru[MeSH] OR philippines[MeSH] OR puerto rico[MeSH] OR russia[MeSH] OR sao tome and principe[MeSH] OR senegal[MeSH] OR sierra leone[MeSH] OR Solomon islands[MeSH] OR somalia[MeSH] OR south sudan[MeSH] OR sri lanka[MeSH] OR sudan[MeSH] OR syria[MeSH] OR tajikistan[MeSH] OR tanzania[MeSH] OR timor-leste[MeSH] OR togo[MeSH] OR tonga[MeSH] OR turkey[MeSH] OR Tuvalu[MeSH] OR uganda[MeSH] OR ukraine[MeSH] OR uzbekistan[MeSH] OR vanuatu[MeSH] OR venezuela[MeSH] OR west bank and gaza[MeSH] OR west timor[MeSH] OR yemen[MeSH] OR zambia[MeSH] OR zimbabwe[MeSH])) OR (((afghanistan[Title/Abstract] OR angola[Title/Abstract] OR armenia[Title/Abstract] OR azerbaijan[Title/Abstract] OR bangladesh[Title/Abstract] OR benin[Title/Abstract] OR bolivia[Title/Abstract] OR bosnia and herzegovina[Title/Abstract] OR burkina faso[Title/Abstract] OR burundi[Title/Abstract] OR cambodia[Title/Abstract] OR cameroon[Title/Abstract] OR central african republic[Title/Abstract] OR chad[Title/Abstract] OR Chechnya[Title/Abstract] OR chile[Title/Abstract] OR colombia[Title/Abstract] OR comoros[Title/Abstract] OR democratic republic of the congo[Title/Abstract] OR congo[Title/Abstract] OR cote d’Ivoire[Title/Abstract] OR cuba[Title/Abstract] OR djibouti[Title/Abstract] OR east timor[Title/Abstract] OR ecuador[Title/Abstract] OR el salvador[Title/Abstract] OR eritrea[Title/Abstract] OR eswatini[Title/Abstract] OR ethiopia[Title/Abstract] OR fiji[Title/Abstract] OR gambia[Title/Abstract] OR grenada[Title/Abstract] OR guatemala[Title/Abstract] OR guinea[Title/Abstract] OR guinea-bissau[Title/Abstract] OR guyana[Title/Abstract] OR haiti[Title/Abstract] OR honduras[Title/Abstract] OR indonesia[Title/Abstract] OR iran[Title/Abstract] OR iraq[Title/Abstract] OR jordan[Title/Abstract] OR kenya[Title/Abstract] OR Kiribati[Title/Abstract] OR democratic people’s republic of korea[Title/Abstract] OR republic of korea[Title/Abstract] OR kosovo[Title/Abstract] OR kyrgyzstan[Title/Abstract] OR laos[Title/Abstract] OR lebanon[Title/Abstract] OR lesotho[Title/Abstract] OR liberia[Title/Abstract] OR libya[Title/Abstract] OR republic of north macedonia[Title/Abstract] OR madagascar[Title/Abstract] OR malawi[Title/Abstract] OR mali[Title/Abstract] OR micronesia[Title/Abstract] OR mauritania[Title/Abstract] OR mozambique[Title/Abstract] OR myanmar[Title/Abstract] OR namibia[Title/Abstract] OR nepal[Title/Abstract] OR netherlands antilles[Title/Abstract] OR nicaragua[Title/Abstract] OR niger[Title/Abstract] OR nigeria[Title/Abstract] OR occupied Palestinian territory[Title/Abstract] OR pakistan[Title/Abstract] OR papua new guinea[Title/Abstract] OR peru[Title/Abstract] OR philippines[Title/Abstract] OR puerto rico[Title/Abstract] OR russia[Title/Abstract] OR sao tome and principe[Title/Abstract] OR senegal[Title/Abstract] OR sierra leone[Title/Abstract] OR Solomon islands[Title/Abstract] OR somalia[Title/Abstract] OR south sudan[Title/Abstract] OR sri lanka[Title/Abstract] OR sudan[Title/Abstract] OR syria[Title/Abstract] OR tajikistan[Title/Abstract] OR tanzania[Title/Abstract] OR timor-leste[Title/Abstract] OR togo[Title/Abstract] OR tonga[Title/Abstract] OR turkey[Title/Abstract] OR Tuvalu[Title/Abstract] OR uganda[Title/Abstract] OR ukraine[Title/Abstract] OR uzbekistan[Title/Abstract] OR vanuatu[Title/Abstract] OR venezuela[Title/Abstract] OR west bank and gaza[Title/Abstract] OR west timor[Title/Abstract] OR yemen[Title/Abstract] OR zambia[Title/Abstract] OR zimbabwe[Title/Abstract])) AND ((humanitarian development[MeSH] OR humanitarian development peace[MeSH] OR humanitarian development peace nexus [MeSH] OR double nexus[MeSH] OR triple nexus[MeSH] OR disaster relief [MeSH] OR disaster risk reduction [MeSH] OR disaster-affected[MeSH] OR humanitarian development divide OR humanitarian development continuum[MeSH] OR humanitarian development gap[MeSH] OR relief to development [MeSH] OR linking relief, rehabilitation and development[MeSH] OR fragile state[MeSH] OR fragility[MeSH] OR fragile setting[MeSH] or fragile country[MeSH] OR conflict-affected[MeSH] OR war-torn[MeSH] OR war-affected[MeSH] OR displacement setting[MeSH] OR refugee setting[MeSH] OR emergency relief [MeSH] OR complex emergency [MeSH] OR protracted crises[MeSH]) OR (humanitarian development[Title/Abstract] OR humanitarian development peace[Title/Abstract] OR humanitarian development peace nexus [Title/Abstract] OR double nexus[Title/Abstract] OR triple nexus[Title/Abstract] OR disaster relief [Title/Abstract] OR disaster risk reduction [Title/Abstract] OR disaster-affected[Title/Abstract] OR humanitarian development divide OR humanitarian development continuum[Title/Abstract] OR humanitarian development gap[Title/Abstract] OR relief to development [Title/Abstract] OR linking relief, rehabilitation and development[Title/Abstract] OR fragile state[Title/Abstract] OR fragility[Title/Abstract] OR fragile setting[Title/Abstract] or fragile country[Title/Abstract] OR conflict-affected[Title/Abstract] OR war-torn[Title/Abstract] OR war-affected[Title/Abstract] OR displacement setting[Title/Abstract] OR refugee setting[Title/Abstract] OR emergency relief [Title/Abstract] OR complex emergency [Title/Abstract] OR protracted crises[Title/Abstract])) AND (((Maternal health[MeSH] OR child health[MeSH] OR infant health[MeSH] OR immunization[MeSH] OR vaccines[MeSH] OR vaccination[MeSH] OR mass vaccination[MeSH] OR sexual health[MeSH] OR adolescent health[MeSH] OR family planning services[MeSH] OR gynecology[MeSH] OR contraception[MeSH] OR birth intervals[MeSH] OR adolescent health[MeSH])) OR (Maternal health[Title/Abstract] OR child health[Title/Abstract] OR maternal child health[Title/Abstract] OR maternal and child health[Title/Abstract] OR newborn health[Title/Abstract] OR neonatal health[Title/Abstract] OR immunizations[Title/Abstract] OR vaccines[Title/Abstract] Or vaccinations[Title/Abstract] or sexual[Title/Abstract] or reproductive[Title/Abstract] OR sexual health[Title/Abstract] OR sexual and reproductive health[Title/Abstract] OR sexual and reproductive health and rights[Title/Abstract] OR sexual and reproductive health rights[Title/Abstract] OR obstetric[Title/Abstract] OR gynecology[Title/Abstract] OR gynecological[Title/Abstract] OR contraception[Title/Abstract] or contraceptives[Title/Abstract] OR family planning[Title/Abstract] OR birth spacing[Title/Abstract] OR “healthy timing and spacing of pregnancy”[Title/Abstract] OR intrapartum[Title/Abstract] OR antenatal[Title/Abstract] OR prenatal[Title/Abstract] OR perinatal[Title/Abstract] OR postnatal[Title/Abstract] OR adolescent health[Title/Abstract]))

**Results: 313 results**

**Scopus Database Search**

- Search Term/Concept 1—LMICs, fragile and conflict-affected states
- Search Term/Concept 2—humanitarian development peace nexus (HDPN)
- Search Term/Concept 3—maternal and child health/sexual and reproductive health (SRMNCAH)

**LMIC Filter (Search Term 1)**

“afghanistan” OR “ angola” OR “ armenia” OR “ azerbaijan” OR “ bangladesh” OR “ benin” OR “ bolivia” OR {bosnia and Herzegovina} OR {burkina faso} OR “ burundi” OR “ cambodia” OR “ cameroon” OR {central african republic} OR “ chad” OR “ Chechnya” OR “ chile” OR “ colombia” OR “ comoros” OR {democratic republic of the congo} OR “ congo” OR {cote d’Ivoire} OR “ cuba” OR “ djibouti” OR {east timor} OR “ ecuador” OR {el Salvador} OR “ eritrea” OR “ eswatini” OR “ ethiopia” OR “ fiji” OR “ gambia” OR “ grenada” OR “ guatemala” OR “ guinea” OR {guinea-bissau} OR “ guyana” OR “ haiti” OR “ honduras” OR “ indonesia” OR “ iran” OR “ iraq” OR “ jordan” OR “ kenya” OR “ Kiribati” OR {democratic people’s republic of korea} OR {republic of korea} OR “ kosovo” OR “ kyrgyzstan” OR “ laos” OR “ lebanon” OR “ lesotho” OR “ liberia” OR “ libya” OR {republic of north Macedonia} OR “ madagascar” OR “ malawi” OR “ mali” OR “ micronesia” OR “ mauritania” OR “ mozambique” OR “ myanmar” OR “ namibia” OR “ nepal” OR {netherlands Antilles} OR “ nicaragua” OR “ niger” OR “ nigeria” OR {occupied Palestinian territory} OR “ pakistan” OR {papua new guinea} OR “ peru” OR “ philippines” OR {puerto rico} OR “ russia” OR {sao tome and principe} OR “ senegal” OR {sierra leone} OR {Solomon islands} OR “ somalia” OR {south sudan} OR {sri lanka} OR “ sudan” OR “ syria” OR “ tajikistan” OR “ tanzania” OR {timor-leste} OR “ togo” OR “ tonga” OR “ turkey” OR “ Tuvalu” OR “ uganda” OR “ ukraine” OR “ uzbekistan” OR “ vanuatu” OR “ venezuela” OR {west bank and gaza} OR {west timor} OR “ yemen” OR “ zambia” OR “ Zimbabwe”

**AND: Humanitarian-Development-Peace Nexus Concepts (Search Term 2)**

{humanitarian development} OR { humanitarian development peace} OR { humanitarian development peace nexus} OR { double nexus} OR { triple nexus} OR { disaster relief} OR { disaster risk reduction} OR { disaster-affected} OR { humanitarian development divide} OR { humanitarian development continuum} OR { humanitarian development gap} OR { relief to development} OR { linking relief, rehabilitation and development} OR { fragile state} OR { fragility} OR { fragile setting} OR { fragile country} OR { conflict-affected} OR { war-torn} OR { war-affected} OR { displacement setting} OR { refugee setting} OR { emergency relief} OR { complex emergency} OR { protracted crises}

**AND: SRMNCAH Interventions (Search Term 3)**

{Maternal health} OR { child health} OR { maternal child health} OR { maternal and child health} OR { newborn health} OR { neonatal health} OR { immunizations} OR { vaccines} OR { vaccinations} OR { sexual} OR { reproductive} OR { sexual health} OR { sexual and reproductive health} OR { sexual and reproductive health and rights} OR { sexual and reproductive health rights} OR { obstetric} OR { gynecology} OR { gynecological} OR { contraception} OR { contraceptives} OR { family planning} OR { birth spacing} OR {healthy timing and spacing of pregnancy} OR { intrapartum} OR { antenatal} OR { prenatal} OR { perinatal} OR { postnatal} OR { adolescent health}

**SEARCH RESULTS: 402**

*Article title, Abstract, Keywords

**Web of Science Database Search**

(AB=({ maternal and child health} OR { newborn health} OR { neonatal health} OR { sexual and reproductive health} OR { sexual and reproductive health and rights} OR{ family planning} OR {healthy timing and spacing of pregnancy} OR { intrapartum} OR { adolescent health}))

(((((((AB=((“afghanistan” OR “ angola” OR “ armenia” OR “ azerbaijan” OR “ bangladesh” OR “ benin” OR “ bolivia” OR {bosnia and Herzegovina} OR {burkina faso} OR “ burundi” OR “ cambodia” OR “ cameroon” OR {central african republic} OR “ chad” OR “ Chechnya” OR “ chile” OR “ colombia” OR “ comoros” OR {democratic republic of the congo} OR “ congo” OR {cote d’Ivoire} OR “ cuba” OR “ djibouti” OR {east timor} OR “ ecuador” OR {el Salvador} OR “ eritrea” OR “ eswatini”))) OR AB=((“ ethiopia” OR “ fiji” OR “ gambia” OR “ grenada” OR “ guatemala” OR “ guinea” OR {guinea-bissau} OR “ guyana” OR “ haiti” OR “ honduras” OR “ indonesia” OR “ iran” OR “ iraq” OR “ jordan” OR “ kenya” OR “ Kiribati” OR {democratic people’s republic of korea} OR {republic of korea} OR “ kosovo” OR “ kyrgyzstan” OR “ laos” OR “ lebanon” OR “ lesotho” OR “ liberia” OR “ libya” OR {republic of north Macedonia} OR “ madagascar” OR “ malawi” OR “ mali” OR “ micronesia”))) OR AB=((“ mauritania” OR “ mozambique” OR “ myanmar” OR “ namibia” OR “ nepal” OR {netherlands Antilles} OR “ nicaragua” OR “ niger” OR “ nigeria” OR {occupied Palestinian territory} OR “ pakistan” OR {papua new guinea} OR “ peru” OR “ philippines” OR {puerto rico} OR “ russia” OR {sao tome and principe} OR “ senegal” OR {sierra leone} OR {Solomon islands} OR “ somalia” OR {south sudan}))) OR AB=(({sri lanka} OR “ sudan” OR “ syria” OR “ tajikistan” OR “ tanzania” OR {timor-leste} OR “ togo” OR “ tonga” OR “ turkey” OR “ Tuvalu” OR “ uganda” OR “ ukraine” OR “ uzbekistan” OR “ vanuatu” OR “ venezuela” OR {west bank and gaza} OR {west timor} OR “ yemen” OR “ zambia” OR “ Zimbabwe”))) AND (AB=({humanitarian development} OR { humanitarian development peace} OR { humanitarian development peace nexus} OR { double nexus} OR { triple nexus} OR { disaster relief} OR { disaster risk reduction} OR { disaster-affected} OR { humanitarian development divide} OR { humanitarian development continuum} OR { humanitarian development gap} OR { relief to development} )) OR AB=({ linking relief, rehabilitation and development} OR { fragile state} OR { fragility} OR { fragile setting} OR { fragile country} OR { conflict-affected} OR { war-torn} OR { war-affected} OR { displacement setting} OR { refugee setting} OR { emergency relief} OR { complex emergency} OR { protracted crises}))) AND (AB=({ maternal and child health} OR { newborn health} OR { neonatal health} OR { sexual and reproductive health} OR { sexual and reproductive health and rights} OR{ family planning} OR {healthy timing and spacing of pregnancy} OR { intrapartum} OR { adolescent health})))

**Results: 781**
