## Supplementary material for "The Humanitarian-Development Nexus and Sexual and Reproductive Health Interventions in Fragile Settings: A Scoping Review": S3 File.docx

**S3 File. Grey Literature Sources**

- ReliefWeb

- HumanitarianResponse.info

- Inter-Agency Working Group on Reproductive Health in Humanitarian Crises (IAWG)

- USAID Development Experience Clearinghouse (DEC)

- USAID Nutrition Resource Hub

- Healthy Newborn Network

- Child Health Task Force

- FP2030

- CORE Group

- Implementing Best Practices (IBP) Network

- Maternal and Child Survivor Program (MCSP)

- Maternal and Child Health Integrated Program (MCHIP)

- Knowledge Success

- Inter-Agency Standing Committee (IASC)

- The World Bank

- United Nations websites: WHO, UNFPA, OCHA, UNICEF, UNHCR
