## Supplementary material for "The Humanitarian-Development Nexus and Sexual and Reproductive Health Interventions in Fragile Settings: A Scoping Review": S4 File.docx

| **S4 File. Low- and Middle-Income Countries by Fragility Inclusion Criteria** | | | | | | | |
| --- | --- | --- | --- | --- | --- | --- | --- |
|  |  |  | **Fragility Designation** | | | | |
| **No.** | **Country** | **LMIC Designation** | **Low Income Category Under Stress**  **(LICUS)** | **Fragile State** | **Fragile Situation** | **Fragile and Conflict-Affected Situation (FCS)** | **Humanitarian Response Plan (HRP)/**  **Flash Appeal** |
| 1 | Afghanistan | Low Income | 🗸 | 🗸 | 🗸 | 🗸 | 🗸 |
| 2 | Angola | Lower middle income | 🗸 |  | 🗸 |  | 🗸 |
| 3 | Armenia | Upper middle income |  | 🗸 |  |  |  |
| 4 | Azerbaijan | Upper middle income |  | 🗸 |  |  |  |
| 5 | Bangladesh | Lower middle income |  |  |  |  | 🗸 |
| 6 | Benin | Lower middle income |  |  |  |  | 🗸 |
| 7 | Bolivia | Lower middle income |  |  |  |  | 🗸 |
| 8 | Bosnia and Herzegovina | Upper middle income |  |  |  |  |  |
| 9 | Burkina Faso | Low income |  | 🗸 |  | 🗸 | 🗸 |
| 10 | Burundi | Low income | 🗸 | 🗸 | 🗸 | 🗸 | 🗸 |
| 11 | Cambodia | Lower middle income | 🗸 |  |  |  |  |
| 12 | Cameroon | Lower middle income | 🗸 | 🗸 |  | 🗸 | 🗸 |
| 13 | Central African Republic | Low income | 🗸 | 🗸 | 🗸 | 🗸 | 🗸 |
| 14 | Chad | Low income | 🗸 | 🗸 | 🗸 | 🗸 | 🗸 |
| 15 | Chechnya | N/A, Territory |  |  |  |  | 🗸 |
| 16 | Colombia | Upper middle income |  |  |  |  | 🗸 |
| 17 | Comoros | Lower middle income | 🗸 | 🗸 | 🗸 | 🗸 |  |
| 18 | Congo, Democratic Republic | Low income | 🗸 | 🗸 | 🗸 | 🗸 | 🗸 |
| 19 | Congo, Republic | Lower middle income | 🗸 | 🗸 | 🗸 | 🗸 | 🗸 |
| 20 | Côte d’Ivoire | Lower middle income | 🗸 |  | 🗸 |  | 🗸 |
| 21 | Cuba | Upper middle income |  |  |  |  | 🗸 |
| 22 | Djibouti | Lower middle income | 🗸 |  | 🗸 |  | 🗸 |
| 23 | East Timor | N/A, Territory |  |  |  |  | 🗸 |
| 24 | Ecuador | Upper middle income |  |  |  |  | 🗸 |
| 25 | El Salvador | Lower middle income |  |  |  |  | 🗸 |
| 26 | Eritrea | Low income | 🗸 | 🗸 | 🗸 | 🗸 | 🗸 |
| 27 | Eswatini (formerly Swaziland) | Lower middle income |  |  |  |  | 🗸 |
| 28 | Ethiopia | Low income |  | 🗸 |  | 🗸 | 🗸 |
| 29 | Fiji | Upper middle income |  |  |  |  | 🗸 |
| 30 | Gambia, The | Low income | 🗸 | 🗸 | 🗸 |  | 🗸 |
| 31 | Grenada | Upper middle income |  |  |  |  | 🗸 |
| 32 | Guatemala | Upper middle income |  |  |  |  | 🗸 |
| 33 | Guinea | Low income | 🗸 |  | 🗸 |  | 🗸 |
| 34 | Guinea-Bissau | Low income | 🗸 | 🗸 | 🗸 | 🗸 | 🗸 |
| 35 | Guyana | Upper middle income |  |  |  |  | 🗸 |
| 36 | Haiti | Lower middle income | 🗸 | 🗸 | 🗸 | 🗸 | 🗸 |
| 37 | Honduras | Lower middle income |  |  |  |  | 🗸 |
| 38 | Indonesia | Lower middle income |  |  |  |  | 🗸 |
| 39 | Iran, Islamic Rep. | Lower middle income |  |  |  |  | 🗸 |
| 40 | Iraq | Upper middle income |  |  |  |  | 🗸 |
| 41 | Jordan | Upper middle income |  |  |  |  | 🗸 |
| 42 | Kenya | Lower middle income |  |  |  |  | 🗸 |
| 43 | Kiribati | Lower middle income | 🗸 | 🗸 | 🗸 |  |  |
| 44 | Korea, Dem. People's Rep. | Low income |  |  |  |  | 🗸 |
| 45 | Kosovo | Upper middle income | 🗸 | 🗸 | 🗸 | 🗸 |  |
| 46 | Kyrgyz Republic | Lower middle income |  |  |  |  | 🗸 |
| 47 | Lao PDR | Lower middle income | 🗸 | 🗸 |  |  | 🗸 |
| 48 | Lebanon | Lower middle income |  | 🗸 | 🗸 | 🗸 | 🗸 |
| 49 | Lesotho | Lower middle income |  |  |  |  | 🗸 |
| 50 | Liberia | Low income | 🗸 | 🗸 | 🗸 |  | 🗸 |
| 51 | Libya | Upper middle income |  | 🗸 | 🗸 | 🗸 | 🗸 |
| 52 | Madagascar | Low income |  |  | 🗸 |  | 🗸 |
| 53 | Malawi | Low income |  |  | 🗸 |  | 🗸 |
| 54 | Mali | Low income |  | 🗸 | 🗸 | 🗸 | 🗸 |
| 55 | Marshall Islands | Upper middle income | 🗸 | 🗸 | 🗸 | 🗸 |  |
| 56 | Mauritania | Lower middle income | 🗸 |  |  |  | 🗸 |
| 57 | Micronesia, Fed. Sts. | Lower middle income |  | 🗸 | 🗸 | 🗸 |  |
| 58 | Mozambique | Low income | 🗸 | 🗸 | 🗸 | 🗸 | 🗸 |
| 59 | Myanmar | Lower middle income |  | 🗸 | 🗸 | 🗸 | 🗸 |
| 60 | Namibia | Upper middle income |  |  |  |  | 🗸 |
| 61 | Nepal | Lower middle income |  |  | 🗸 |  | 🗸 |
| 62 | Nicaragua | Lower middle income |  |  |  |  | 🗸 |
| 63 | Niger | Low income |  | 🗸 |  | 🗸 | 🗸 |
| 64 | Nigeria | Lower middle income | 🗸 | 🗸 |  | 🗸 | 🗸 |
| 65 | Occupied Palestinian Territory | N/A, Territory |  |  |  |  | 🗸 |
| 66 | Pakistan | Lower middle income |  |  |  |  | 🗸 |
| 67 | Papua New Guinea | Lower middle income | 🗸 | 🗸 | 🗸 | 🗸 |  |
| 68 | Peru | Upper middle income |  |  |  |  | 🗸 |
| 69 | Philippines | Lower middle income |  |  |  |  | 🗸 |
| 70 | Russian Federation | Upper middle income |  |  |  |  | 🗸 |
| 71 | São Tomé and Príncipe | Lower middle income | 🗸 |  | 🗸 |  |  |
| 72 | Senegal | Lower middle income |  |  |  |  | 🗸 |
| 73 | Sierra Leone | Low income | 🗸 |  | 🗸 |  | 🗸 |
| 74 | Solomon Islands | Lower middle income | 🗸 | 🗸 | 🗸 | 🗸 | 🗸 |
| 75 | Somalia | Low income | 🗸 | 🗸 | 🗸 | 🗸 | 🗸 |
| 76 | South Sudan | Low income |  | 🗸 | 🗸 | 🗸 | 🗸 |
| 77 | Sri Lanka | Lower middle income |  |  |  |  | 🗸 |
| 78 | Sudan | Low income | 🗸 | 🗸 | 🗸 | 🗸 | 🗸 |
| 79 | Syrian Arab Republic | Low income |  | 🗸 | 🗸 | 🗸 | 🗸 |
| 80 | Tajikistan | Lower middle income | 🗸 |  |  |  | 🗸 |
| 81 | Tanzania | Lower middle income |  |  |  |  | 🗸 |
| 82 | Timor-Leste | Lower middle income | 🗸 | 🗸 | 🗸 | 🗸 | 🗸 |
| 83 | Togo | Low income | 🗸 |  | 🗸 |  | 🗸 |
| 84 | Tonga | Upper middle income | 🗸 |  |  |  |  |
| 85 | Türkiye | Upper middle income |  |  |  |  |  |
| 86 | Tuvalu | Upper middle income |  | 🗸 | 🗸 | 🗸 |  |
| 87 | Uganda | Low income |  |  |  |  | 🗸 |
| 88 | Ukraine | Lower middle income |  |  |  |  | 🗸 |
| 89 | Uzbekistan | Lower middle income | 🗸 |  |  |  |  |
| 90 | Vanuatu | Lower middle income | 🗸 |  |  |  |  |
| 91 | Venezuela, RB | **Not categorized in WB |  | 🗸 |  | 🗸 | 🗸 |
| 92 | West Bank and Gaza | Lower middle income | 🗸 | 🗸 | 🗸 | 🗸 |  |
| 93 | West Timor | N/A, Territory |  |  |  |  | 🗸 |
| 94 | Yemen | Low income | 🗸 | 🗸 | 🗸 | 🗸 | 🗸 |
| 95 | Zambia | Low income |  |  |  |  | 🗸 |
| 96 | Zimbabwe | Lower middle income | 🗸 | 🗸 | 🗸 | 🗸 | 🗸 |
